# Critical Care Resources and Disaster Preparedness Survey 2020

**DOI:** 10.1101/2021.04.08.21254902

**Authors:** Simon Finfer, Naomi Hammond, Bharath Kumar Tirupakuzhi Vijayaraghavan, Lowell Ling, Louise Thwaites, Brett Abbenbroek

## Abstract

**Aim:** To investigate critical care resourcing and the clinical management of sepsis in lower-middle income, upper-middle income and high income countries across the Asia Pacific region.

**Background:** Sepsis is a time-critical complex condition that requires evidence-based care delivered by appropriate levels of well trained, qualified and experienced staff supported by proactive organisational and quality processes, sophisticated technologies and reliable infrastructure. In 2017, the estimated sepsis incidence in the Asia Pacific region ranged from 120 to 200 per 100,000 population in Australia and New Zealand to 2500 to 3400 per 100,000 population in India. Currently, there is limited information on the organisational structures, human resources, clinical standards, laboratory support and the therapeutic options available in the Asia Pacific region to treat sepsis.

**Method:** Prospective electronic survey.

**Results:** Representatives of 59 hospitals from 15 countries responded. Provision of critical care and the management of sepsis varied considerably between lower-middle income, upper-middle income and high income countries. Specific differences include nurse to patient ratios and availability of allied health services.

Conventional organ support modalities such as mechanical ventilation and non-invasive ventilation were commonly available. Even advanced life support like extracorporeal membrane oxygenation was available in at least 60% of surveyed ICUs. However, in contrast, essential monitoring devices including EtCO2 were not universally available.

Lower-middle income countries had considerably lower provisions for isolation and surge capacity to support pandemic and disaster management, though basic personal protective equipment was widely available.

A majority of ICUs used the Surviving Sepsis Campaign guidelines or the adapted version for lower-middle income countries, though only 21% of ICUs in lower-middle income countries used the adapted version. While essential antimicrobials were accessible across most ICUs, availability of reserve antibiotics was limited.

**Conclusion:** The disparities identified in this survey inform healthcare workers and health services, policy makers and governments on the priorities for action to improve the delivery of critical care and sepsis outcomes in this region.

## Introduction

Sepsis is a time-critical complex condition that requires evidence-based care delivered by appropriate levels of well trained, qualified and experienced staff supported by proactive organisational and quality processes, sophisticated technologies and reliable infrastructure. In 2017, the estimated sepsis incidence in the Asia Pacific region ranged from 120 to 200 per 100,000 population in Australia to 2500 to 3400 per 100,000 population in India.^1^

Currently, there is limited information on the organisational structures, human resources, clinical standards, laboratory support, and therapeutic options available in the Asia Pacific region to treat sepsis.^2^ The Asia Pacific Sepsis Alliance (APSA), a regional network of the Global Sepsis Alliance (GSA), conducted a survey across the Asia Pacific including lower middle income (LMIC), upper middle income (UMIC) and high income countries (HIC) to better understand differences in critical care resources.^3^ The purpose of the survey is to inform healthcare workers, services, policy and governments, and facilitate improvements in sepsis care. The survey was conducted during the early phase of the COVID-19 pandemic, so included disaster management and questions specific to COVID-19 patients.

## Method

### Survey design

The electronic survey was adapted from a similar critical care resources survey designed by the Latin America Intensive Care Network^4^, and modified to suit regional needs. Survey development was iterative and was led by a working group of critical care clinicians and researchers from the region. The working group included representatives from both HICs and LMICs. Survey responses were predominantly limited to ‘yes’ or ‘no’ type answers with select questions requiring a quantitative response or a descriptive response. No free text responses were allowed. The draft survey was tested within the survey development group to ensure clarity, logical flow and timeliness. A participant information sheet administered along with the survey provided details of the aims of the survey, instructions for completion and information on consent. The survey was approved Chinese University of Hong Kong Survey and Behavioural Research Ethics (SBRE-19-565).

### Survey administration

The survey was conducted between April 15, 2020 and June 1, 2020. Participants (frontline healthcare workers) were recruited by snowball sampling, first through the APSA network in each country and then through their contacts. Participants were invited by email to complete an online survey. Each respondent provided confirmation that they understood participation was voluntary and that survey completion implied consent for researchers to share and publish the data. The survey was administered using a commercial application-Survey Monkey (SurveyMonkey Inc. San Mateo, California, USA www.surveymonkey.com) and completion time ranged from 7 to 9 minutes. All deidentified survey data was stored on a secure server hosted by The Chinese University of Hong Kong.

## Results

Survey results are presented in figures and the tabulated data provided in Appendix 1.

### Respondents

Representatives of 59 hospitals from 15 countries responded (Figure 1) including 33 LMICs, eight UMICs and 18 HICs.

**Figure 1.**
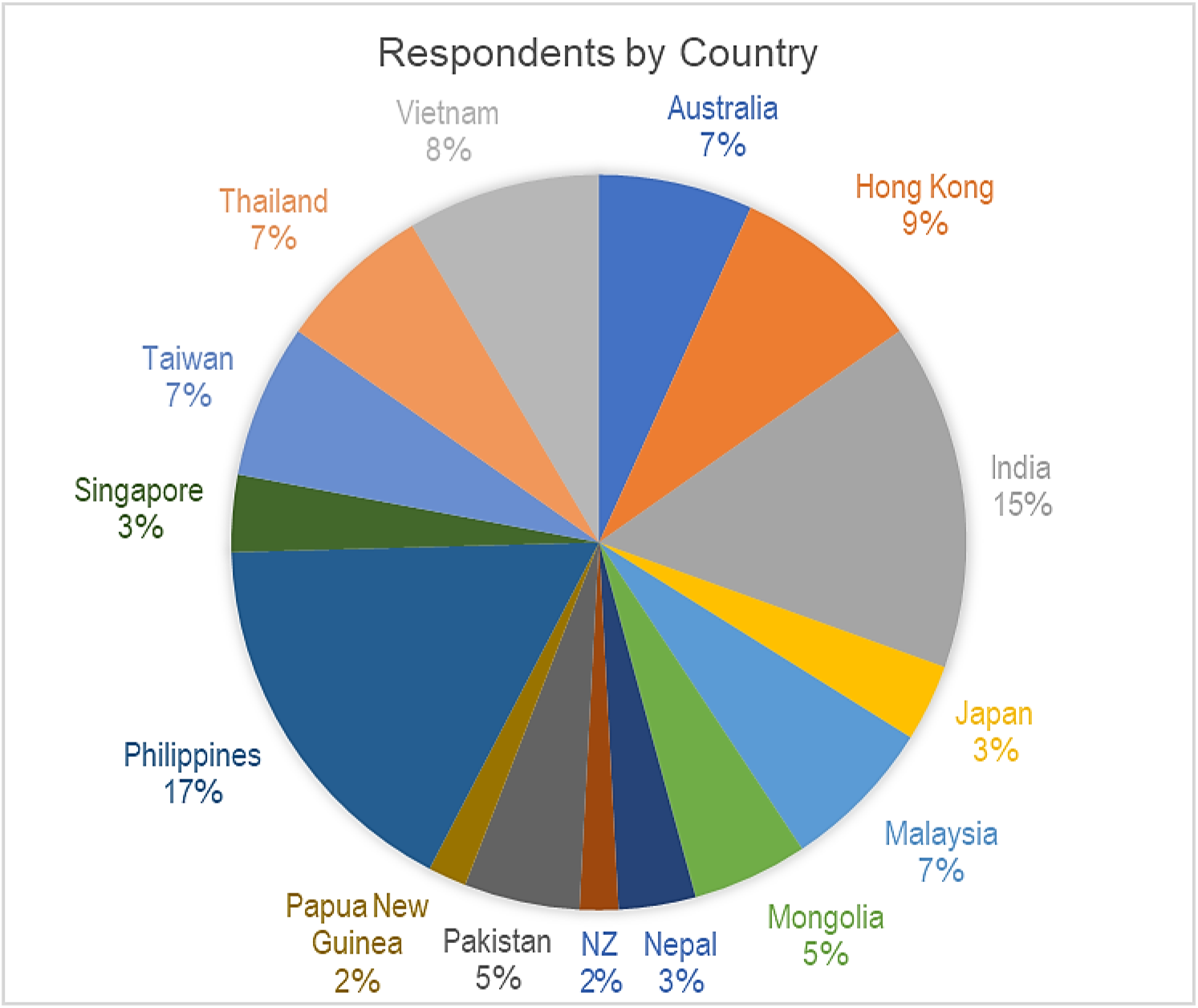
Participating ICUs according to country

The majority of hospitals were tertiary or university hospitals with ICUs capable of treating both adult and paediatric patients with level III facilities (Figure 2).

**Figure 2.**
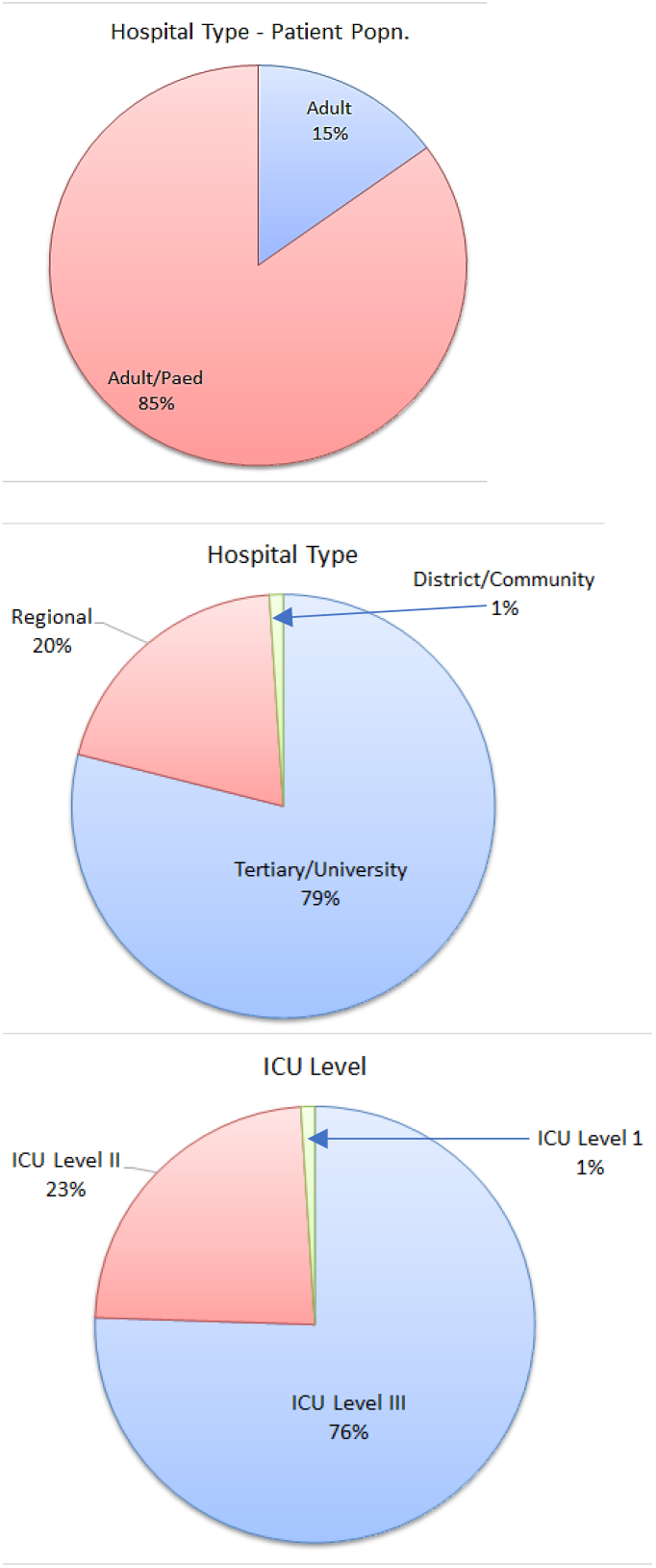
Hospital and ICU type (level)

A level I ICU is capable of providing oxygen, non-invasive monitoring, and more intensive nursing care than on a ward, whereas a level II ICU can provide invasive monitoring and basic life support for a short period.

A level III ICU provides a full spectrum of monitoring and life support technologies, serves as a regional resource for the care of critically ill patients, and may play an active role in developing the specialty of intensive care through research and education.^5^

The majority were large hospitals (median 798 beds, range 50-2000) with a median of 37 (range 3-200) ICU beds and a median of 3000 (range 432-10,000) admissions annually.

Organisational attributes of the hospitals and ICUs are shown in Appendix 1, Table 1 by income group.

**Table 1.** Hospital and ICU characteristics per income group

| Hospital | All |  |  |  | High Income |  |  |  | Upper Middle Income |  |  |  | Lower Middle Income |  |  |  |
| --- | --- | --- | --- | --- | --- | --- | --- | --- | --- | --- | --- | --- | --- | --- | --- | --- |
|  | N<sup>1</sup> | Count | Median | % or (range) | N | Count | Median | % or (range) | N | Count | Median | % or (range) | N | Count | Median | % or (range) |
| Adult |  | 9 |  | 15 |  | 3 |  | 17 |  | 1 |  | 6 |  | 5 |  | 15 |
| Adult & Paediatric | 59 | 50 |  | 85 | 18 | 15 |  | 83 | 8 | 7 |  | 39 | 33 | 28 |  | 85 |
| Community |  | 3 |  | 1 |  | 0 |  | 0 |  | 0 |  | 0 |  | 3 |  | 1 |
| Regional | 59 | 11 |  | 19 | 18 | 6 |  | 33 | 8 | 1 |  | 13 | 33 | 4 |  | 12 |
| Tertiary |  | 45 |  | 76 |  | 12 |  | 67 |  | 7 |  | 87 |  | 26 |  | 79 |
| ICU level 1 | 59 | 3 |  | 1 | 18 | 0 |  | 0 | 8 | 0 |  | 0 | 33 | 3 |  | 9 |
| ICU level 2 | 59 | 12 |  | 23 | 18 | 1 |  | 6 | 8 | 1 |  | 6 | 33 | 10 |  | 30 |
| ICU level 3 | 59 | 44 |  | 76 | 18 | 17 |  | 94 | 8 | 7 |  | 39 | 33 | 20 |  | 61 |
| Hospital beds | 59 |  | 798 | (50-2000) | 18 |  | 1186 | (304-2000) | 8 |  | 1000 | (122-2000) | 33 |  | 599 | (50-2000) |
| ICU beds | 59 |  | 37 | (3-200) | 18 |  | 50 | (15-200) | 8 |  | 49 | (10-170) | 33 |  | 35 | (3-103) |
| HDU beds | 59 |  | 25 | (0-200) | 18 |  | 22 | (0-102) | 8 |  | 35 | (0-66) | 33 |  | 20 | (0-200) |
| Annual ICU admissions | 49 |  | 3000 | (432-10000) | 15 |  | 3510 | (1138-10000) | 7 |  | 3429 | (1005-10000) | 27 |  | 2495 | (432-10000) |
| Day shift<br>Ratio Pt<sup>2</sup> to Dr | 48 |  | 5 | (1-25) | 16 |  | 4 | (1-25) | 6 |  | 4 | (3-10) | 26 |  | 5 | (1-20) |
| Night shift<br>Ratio Pt to Dr | 48 |  | 9.5 | (1-25) | 16 |  | 10 | (1-25) | 6 |  | 8 | (6-25) | 26 |  | 9.5 | (1-30) |
| Day shift<br>Ratio ventilated Pt to Nurse | 48 |  | 2 | (1-8) | 16 |  | 1 | (1-2) | 6 |  | 2 | (1-2) | 26 |  | 2 | (1-8) |
| Night shift<br>Ratio ventilated Pt to Nurse | 48 |  | 2 | (0-8) | 16 |  | 2 | (0-2) | 6 |  | 3 | (1-4) | 26 |  | 2 | (1-8) |
| Day shift<br>Ratio non-vent Pt to Nurse | 48 |  | 2 | (0-50) | 16 |  | 2 | (0-4) | 6 |  | 2 | (1-5) | 26 |  | 2.5 | (1-50) |
| Night shift<br>Ratio non-vent Pt to Nurse | 48 |  | 2 | (0-50) | 16 |  | 2 | (0-4) | 6 |  | 3 | (1-4) | 26 |  | 3 | (1-50) |

1. N = survey responses; 2. Pt = patient

### ICU staffing

Staffing to patient ratios for Doctors and Nurses are summarised in Figure 3.

**Figure 3.**
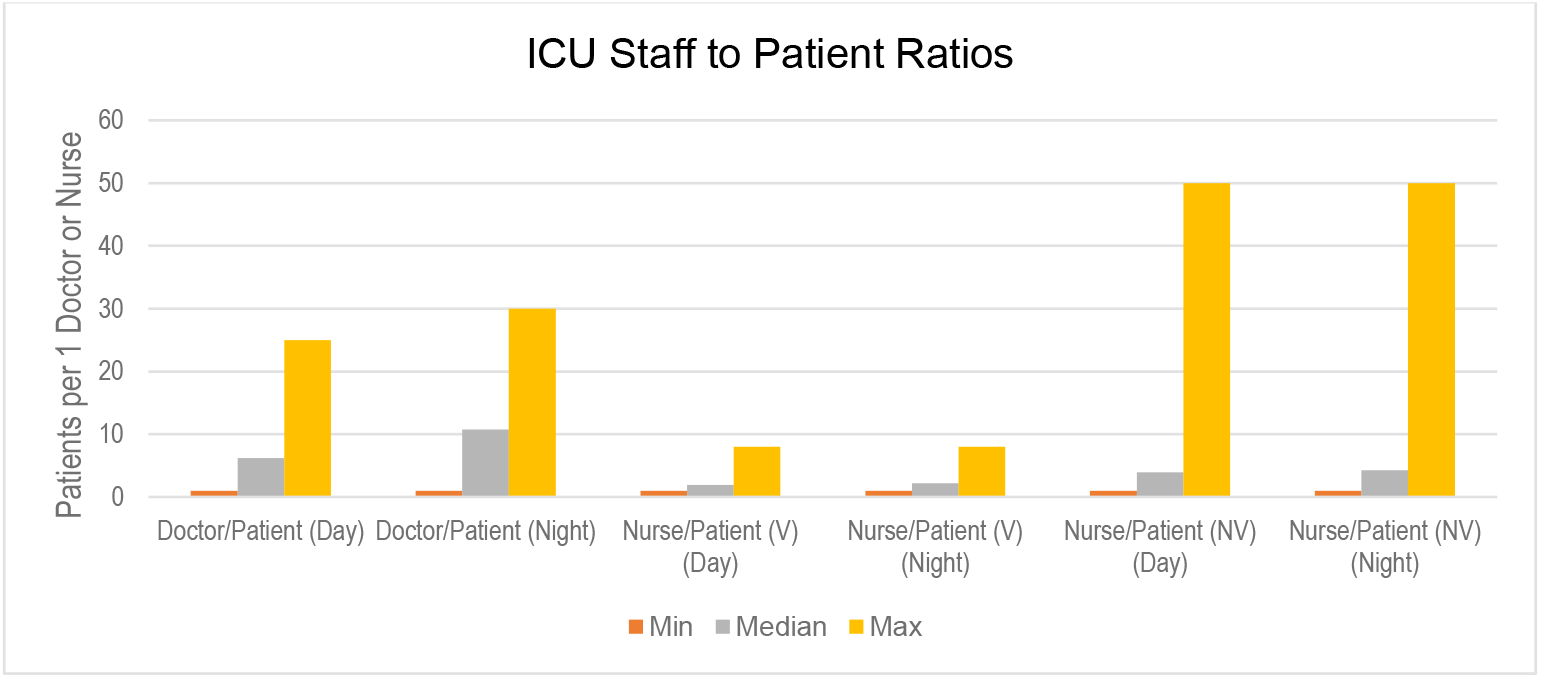
Staff to patient ratios in participating ICUs, Asia Pacific region (NB. V = ventilated, NV = non-ventilated)

Median doctor and nurse to patient ratios are similar across income groups, and all units showed generally reduced staffing numbers at night. Whilst doctor to patient ratios were similar across all income settings, nurse to patient ratios showed a much greater variation, with up to 8 patients per nurse in LMIC ICUs (Table 1).

#### ICU workforce roles

Essential roles including an Intensivist, Physiotherapist, Pharmacist, Nutritionist and Social Worker were available in a majority of ICU’s across the region (Figure 4, Table 1). However, 24/7 access to a physiotherapist was limited in a majority of ICUs in the region.

**Figure 4.**
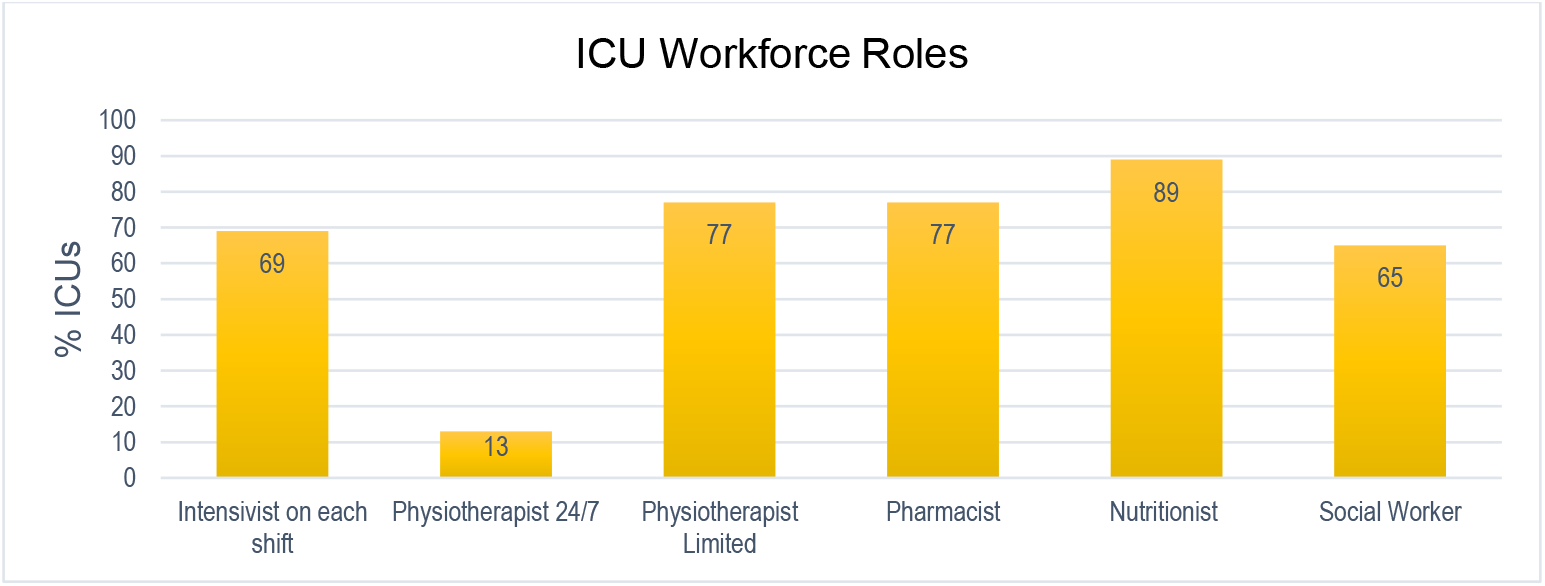
ICU workforce roles, Asia Pacific region

All high and middle income units reported access to physiotherapy (including 24/7 access in 31% of HIC units), while 19% of LMIC reported they had no physiotherapy available in ICU (Figure 5, Table 2).

**Figure 5.**
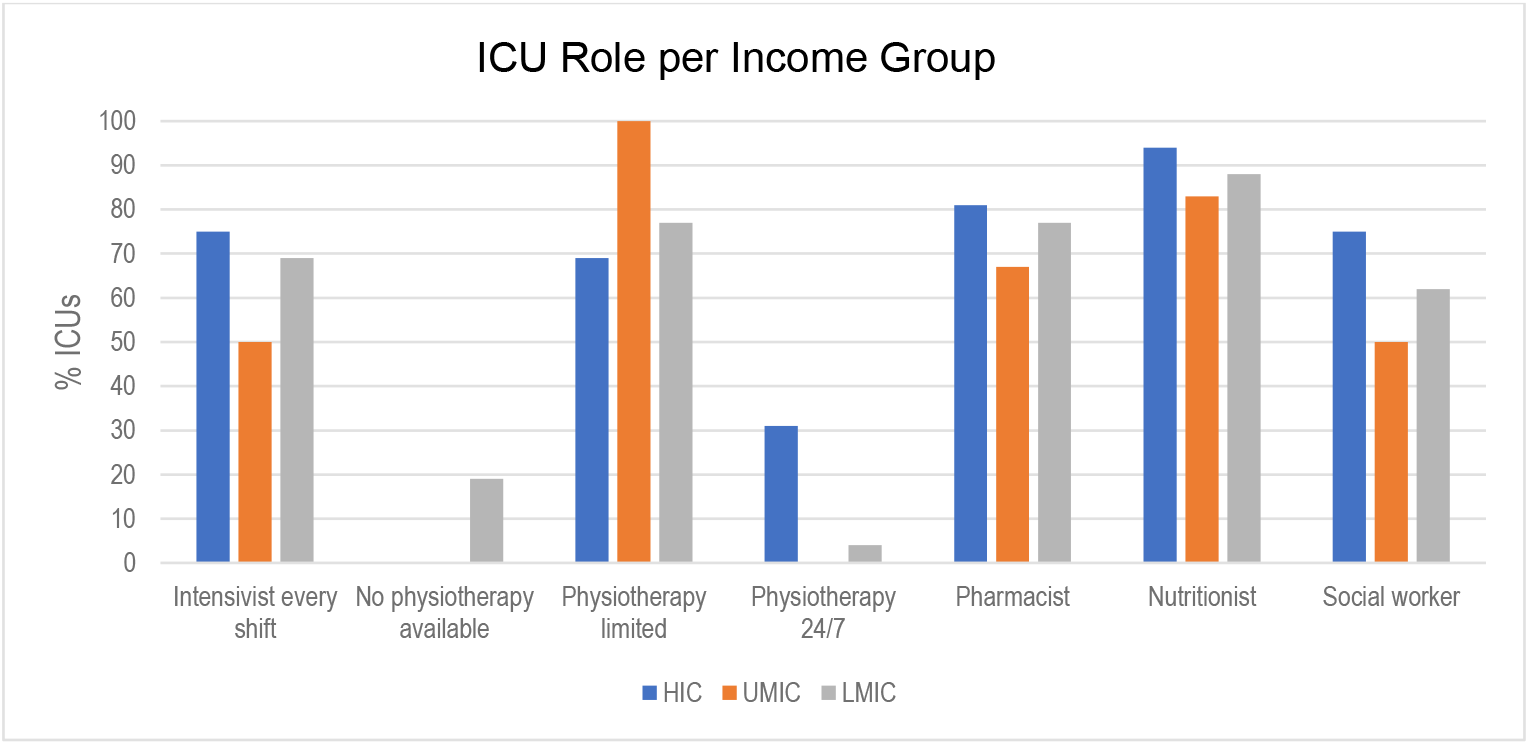
ICU role availability according to country income group

**Table 2.**
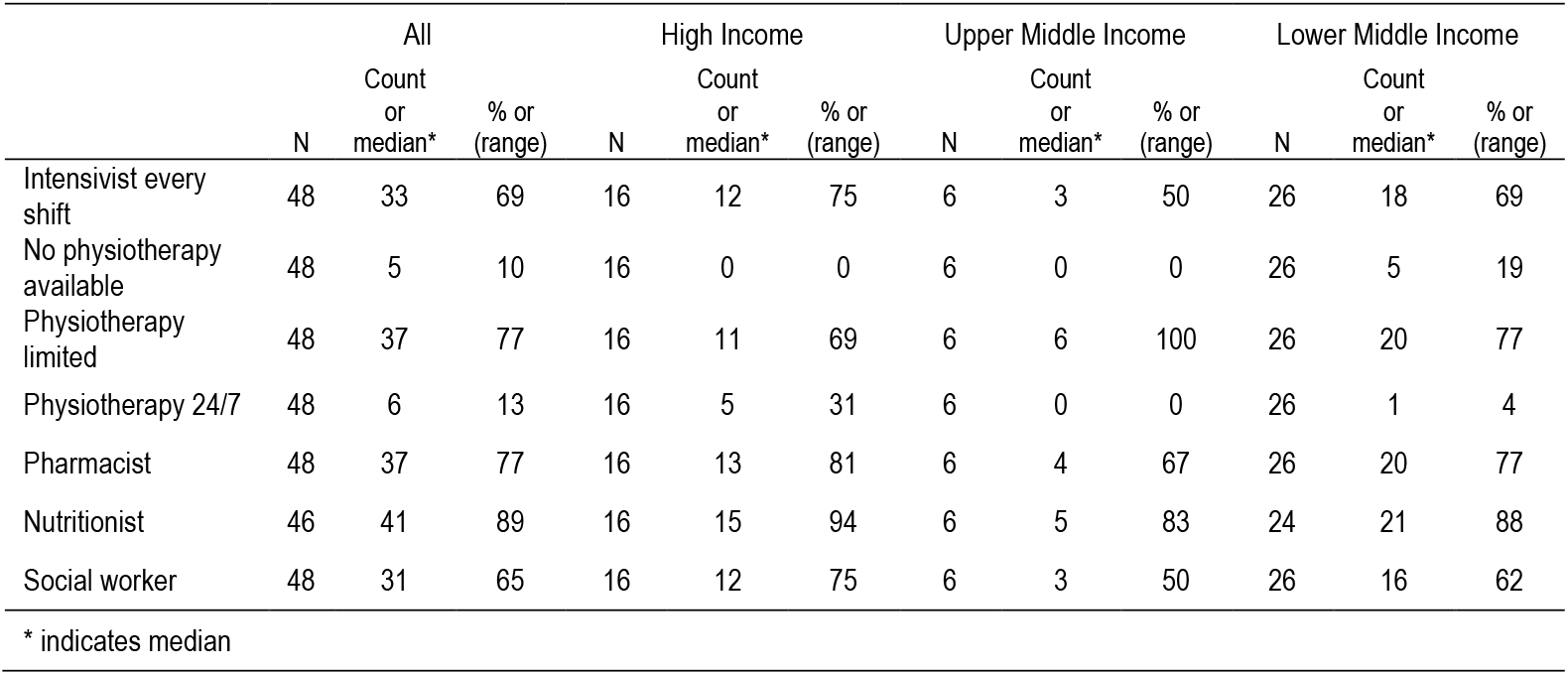
ICU role availability per income group

|  | All |  |  | High Income |  |  | Upper Middle Income |  |  | Lower Middle Income |  |  |
| --- | --- | --- | --- | --- | --- | --- | --- | --- | --- | --- | --- | --- |
|  | N | Count or median* | % or (range) | N | Count or median* | % or (range) | N | Count or median* | % or (range) | N | Count or median* | % or (range) |
| Intensivist every shift | 48 | 33 | 69 | 16 | 12 | 75 | 6 | 3 | 50 | 26 | 18 | 69 |
| No physiotherapy available | 48 | 5 | 10 | 16 | 0 | 0 | 6 | 0 | 0 | 26 | 5 | 19 |
| Physiotherapy limited | 48 | 37 | 77 | 16 | 11 | 69 | 6 | 6 | 100 | 26 | 20 | 77 |
| Physiotherapy 24/7 | 48 | 6 | 13 | 16 | 5 | 31 | 6 | 0 | 0 | 26 | 1 | 4 |
| Pharmacist | 48 | 37 | 77 | 16 | 13 | 81 | 6 | 4 | 67 | 26 | 20 | 77 |
| Nutritionist | 46 | 41 | 89 | 16 | 15 | 94 | 6 | 5 | 83 | 24 | 21 | 88 |
| Social worker | 48 | 31 | 65 | 16 | 12 | 75 | 6 | 3 | 50 | 26 | 16 | 62 |

\* indicates median

### ICU diagnostics

#### Imaging

Standard diagnostic imaging modes were available in all settings (Figure 6, Table 3).

**Figure 6.**
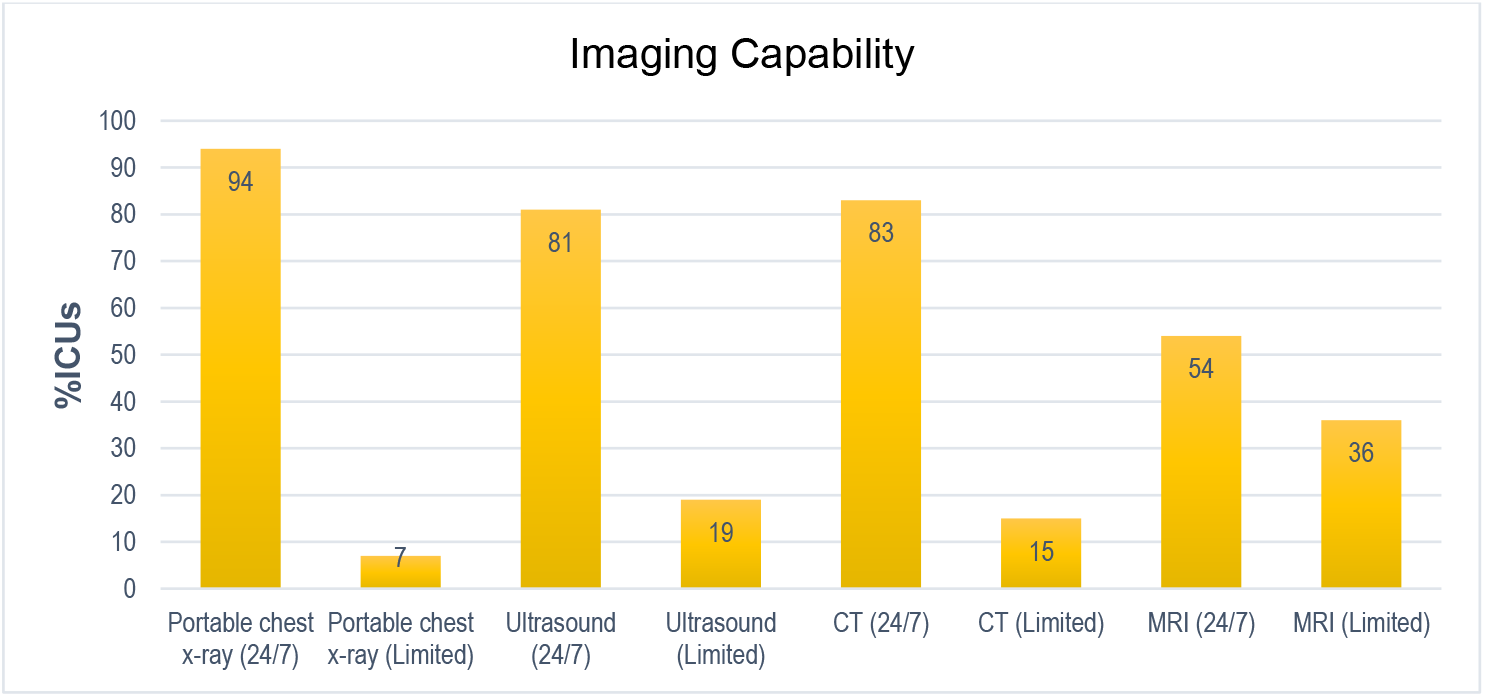
ICU access to different imaging modalities, Asia Pacific region

**Table 3.** ICU diagnostic imaging availability per income group

|  | All |  |  | High Income |  |  | Upper Middle Income |  |  | Lower Middle Income |  |  |
| --- | --- | --- | --- | --- | --- | --- | --- | --- | --- | --- | --- | --- |
|  | N | Count or median* | % or (range) | N | Count or median* | % or (range) | N | Count or median* | % or (range) | N | Count or median* | % or (range) |
| Portable CXR 24/7 | 47 | 44 | 94 | 16 | 14 | 88 | 5 | 5 | 100 | 26 | 25 | 96 |
| Limited portable CXR | 47 | 3 | 6 | 16 | 2 | 12 | 5 | 0 | 0 | 26 | 1 | 4 |
| CT 24/7 | 47 | 39 | 83 | 16 | 13 | 81 | 5 | 5 | 100 | 26 | 21 | 81 |
| Limited CT | 47 | 7 | 15 | 16 | 2 | 12 | 5 | 0 | 0 | 26 | 5 | 19 |
| Ultrasound 24/7 | 47 | 38 | 81 | 16 | 14 | 88 | 5 | 5 | 100 | 26 | 19 | 73 |
| Limited ultrasound | 47 | 9 | 19 | 16 | 2 | 13 | 5 | 0 | 0 | 26 | 7 | 27 |
| MRI 24/7 | 47 | 26 | 55 | 16 | 8 | 50 | 5 | 3 | 60 | 26 | 15 | 58 |
| Limited MRI | 47 | 17 | 36 | 16 | 8 | 50 | 5 | 1 | 20 | 26 | 8 | 31 |
| No MRI | 47 | 4 | 9 | 16 | 0 | 0 | 5 | 1 | 20 | 26 | 3 | 12 |

\* indicates median

Limited availability of diagnostic imaging was particularly evident in lesser resourced LMIC s (Figure 7, Table 3).

**Figure 7.**
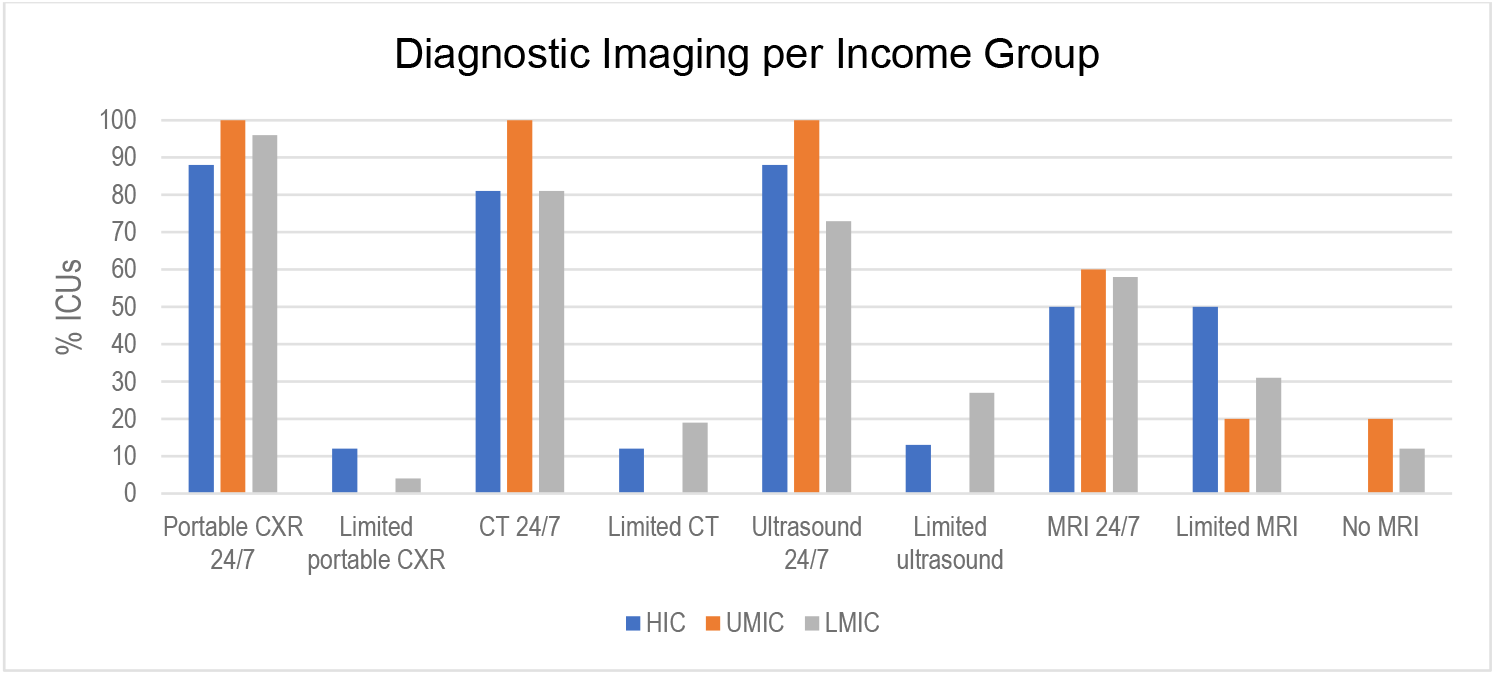
ICU diagnostic imaging availability per income group

#### Pathology

Almost all sites reported 24 hour laboratory availability (92%) and point-of-care testing for arterial blood gases (92%), lactate (68%) and glucose (96%) in the ICU itself (Figure 8, Table 4). Microbiology was also broadly available in ICUs (94%) and offered diagnostic capability for locally relevant pathogens such as dengue and malaria.

**Figure 8.**
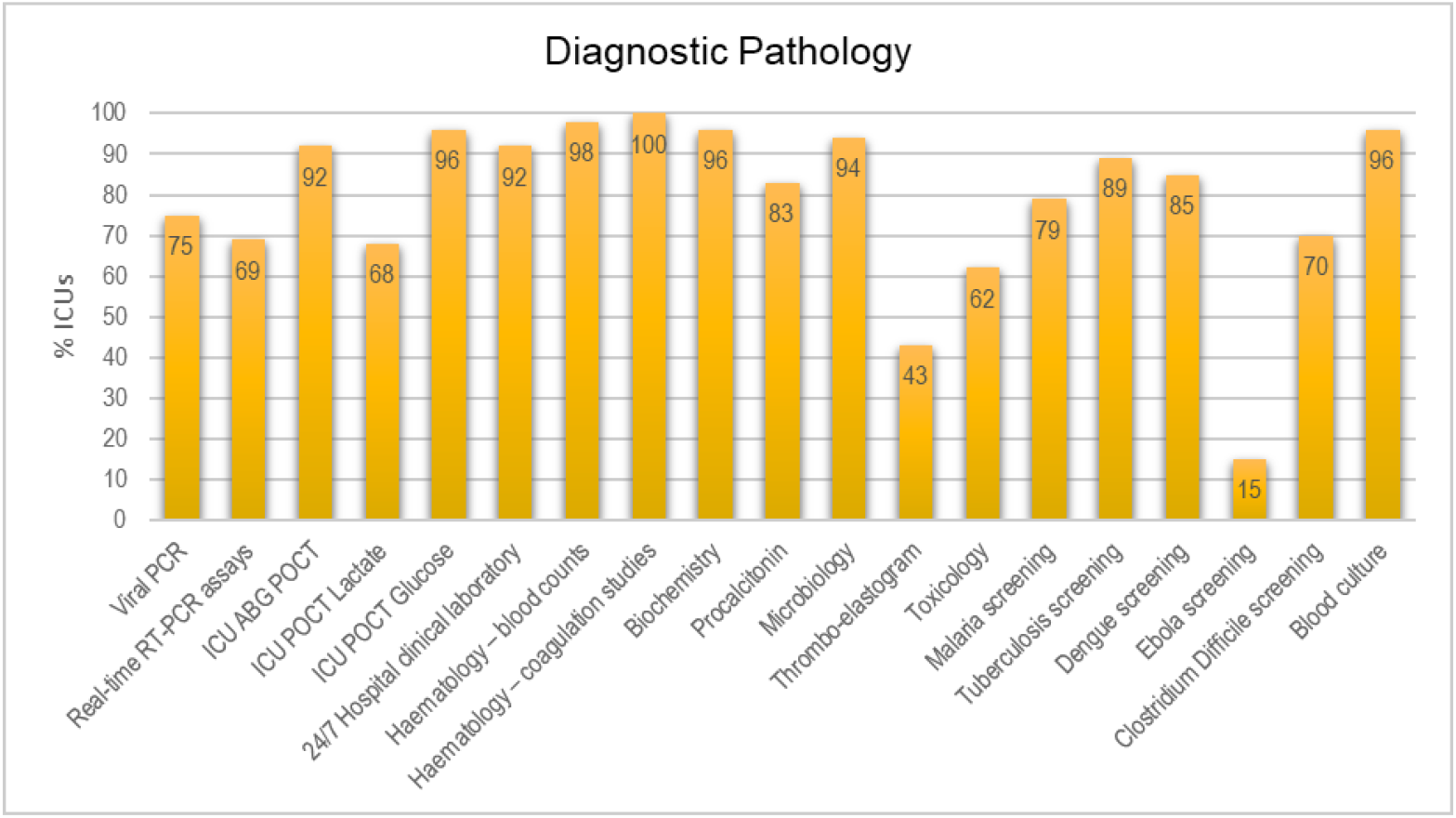
Diagnostic pathology availability in participating ICUs, Asia Pacific region

**Table 4.** ICU diagnostic pathology availability per income group

|  | All |  |  | High Income |  |  | Upper Middle Income |  |  | Lower Middle Income |  |  |
| --- | --- | --- | --- | --- | --- | --- | --- | --- | --- | --- | --- | --- |
|  | N | Count or median* | % or (range) | N | Count or median* | % or (range) | N | Count or median* | % or (range) | N | Count or median* | % or (range) |
| Viral PCR | 47 | 35 | 75 | 16 | 16 | 100 | 5 | 3 | 60 | 26 | 16 | 62 |
| RT PCR SARS-CoV2 | 52 | 36 | 69 | 16 | 12 | 75 | 8 | 7 | 88 | 28 | 17 | 61 |
| Time RT PCR (min) | 51 | 450* | (50-14440) | 15 | 360* | (50-2880) | 8 | 360* | (60-2880) | 28 | 1080* | (240-14440) |
| POC ABG | 47 | 43 | 92 | 16 | 15 | 94 | 5 | 5 | 100 | 26 | 23 | 89 |
| POC Lactate | 47 | 32 | 68 | 16 | 11 | 69 | 5 | 4 | 80 | 26 | 17 | 65 |
| PCO glucose | 47 | 45 | 96 | 16 | 16 | 100 | 5 | 5 | 100 | 26 | 24 | 92 |
| Lab 24/7 | 47 | 44 | 94 | 16 | 15 | 94 | 5 | 5 | 100 | 26 | 24 | 92 |
| Haem-FBC | 47 | 46 | 98 | 16 | 15 | 100 | 5 | 5 | 100 | 26 | 26 | 100 |
| Haem-coagulation | 47 | 47 | 100 | 16 | 16 | 100 | 5 | 5 | 100 | 26 | 26 | 100 |
| Biochemistry | 47 | 45 | 96 | 16 | 15 | 94 | 5 | 5 | 100 | 26 | 25 | 96 |
| Procalcitonin | 46 | 38 | 83 | 16 | 15 | 94 | 4 | 3 | 75 | 26 | 20 | 77 |
| Microbiology | 47 | 44 | 94 | 16 | 14 | 88 | 5 | 5 | 100 | 26 | 25 | 96 |
| Thrombo-elastogram | 47 | 20 | 43 | 16 | 7 | 44 | 5 | 1 | 20 | 26 | 12 | 46 |
| Toxicology | 47 | 29 | 62 | 16 | 12 | 75 | 5 | 4 | 80 | 26 | 13 | 50 |
| Malaria | 47 | 37 | 78 | 16 | 12 | 75 | 5 | 5 | 100 | 26 | 20 | 77 |
| TB | 47 | 42 | 89 | 16 | 14 | 88 | 5 | 5 | 100 | 26 | 23 | 89 |
| Dengue | 47 | 40 | 85 | 16 | 10 | 63 | 5 | 5 | 100 | 26 | 25 | 96 |
| Ebola | 47 | 7 | 15 | 16 | 6 | 38 | 5 | 0 | 0 | 26 | 1 | 96 |
| C.difficile | 47 | 33 | 70 | 16 | 15 | 94 | 5 | 3 | 60 | 26 | 15 | 58 |
| Blood culture | 47 | 45 | 96 | 16 | 15 | 94 | 5 | 5 | 100 | 26 | 25 | 96 |

\* indicates median

Limited access to viral PCR, procalcitonin and C-Difficile testing was evident in UMIC and LMIC ICUs, and less than 50% of LIMIC ICUs could access toxicology services (Figure 9, Table 4). Overall, more UMIC ICU’s had the most types of diagnostic pathology testing available.

**Figure 9.**
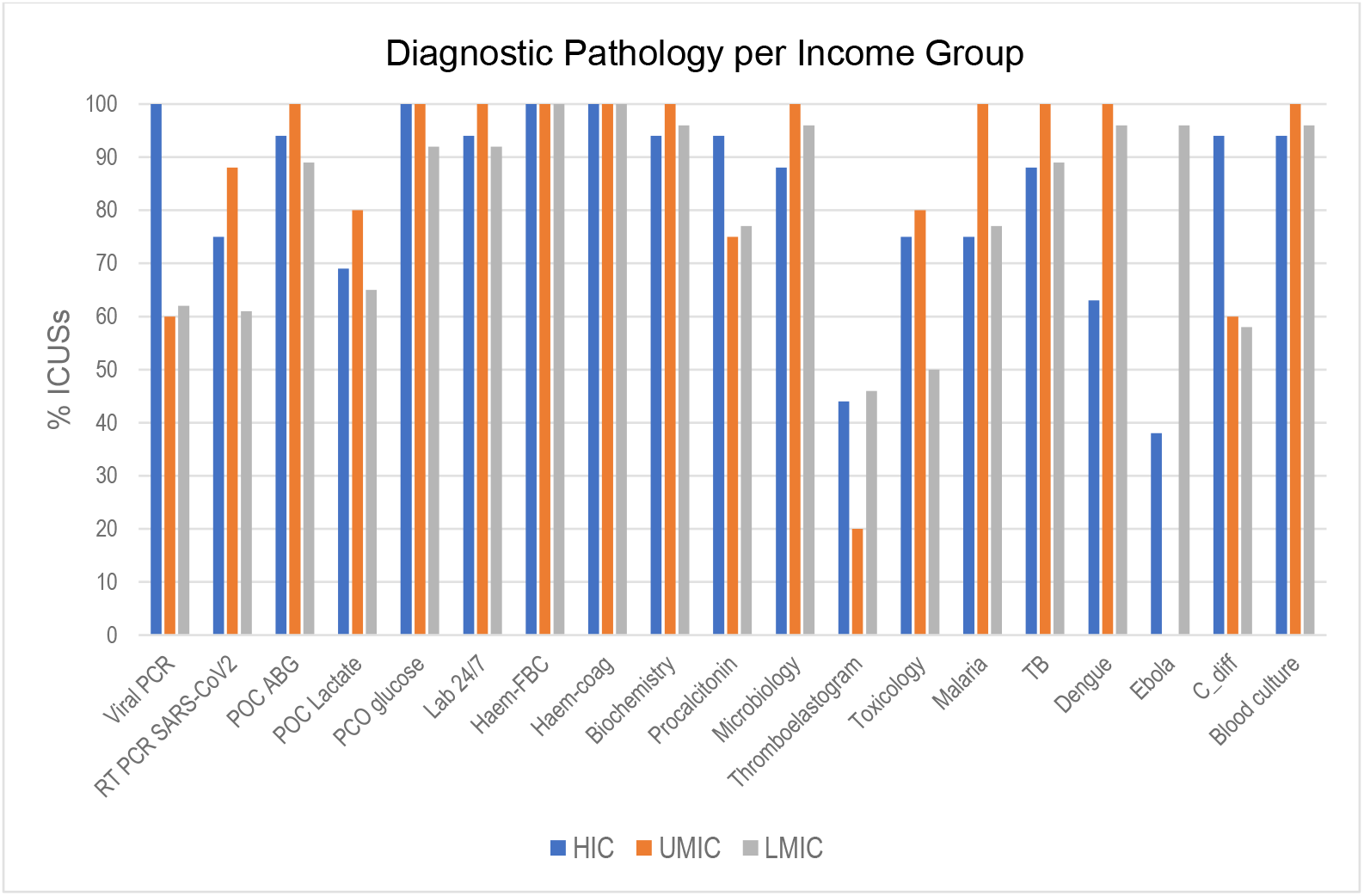
ICU diagnostic pathology availability per income group

The median time for a RT PCR SARS-coV2 test result across the region was 450 min (range 50-14440).

Both HIC and UMIC ICUs reported a shorter median time for a RT PCR SARS-coV2 test result with both income categories taking 360 min, ranging from 50 to 2880 and 60 to 2880 min respectively. In contrast, LMIC ICU’s reported a longer median test result time of 1080 min (range 240-14440).

### ICU clinical care

#### Interventions

Respiratory support modalities were relatively consistent across the region with greater variation in the availability of extracorporeal therapies in ICU including ECMO (62%), CRRT (70%) and plasmapheresis (77%) (Figure 10, Table 5).

**Figure 10.**
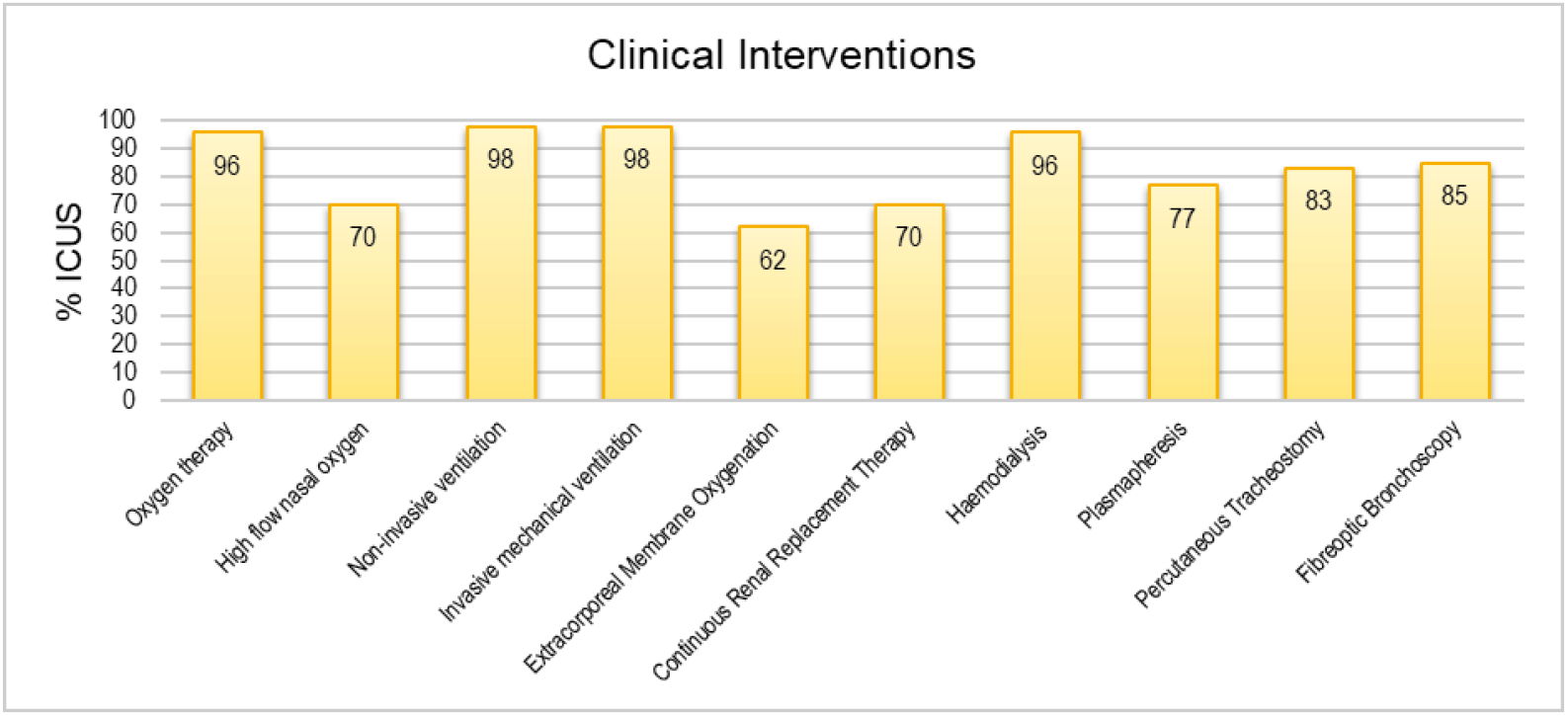
Clinical intervention availability in participating ICUs across the Asia Pacific region

**Table 5.**
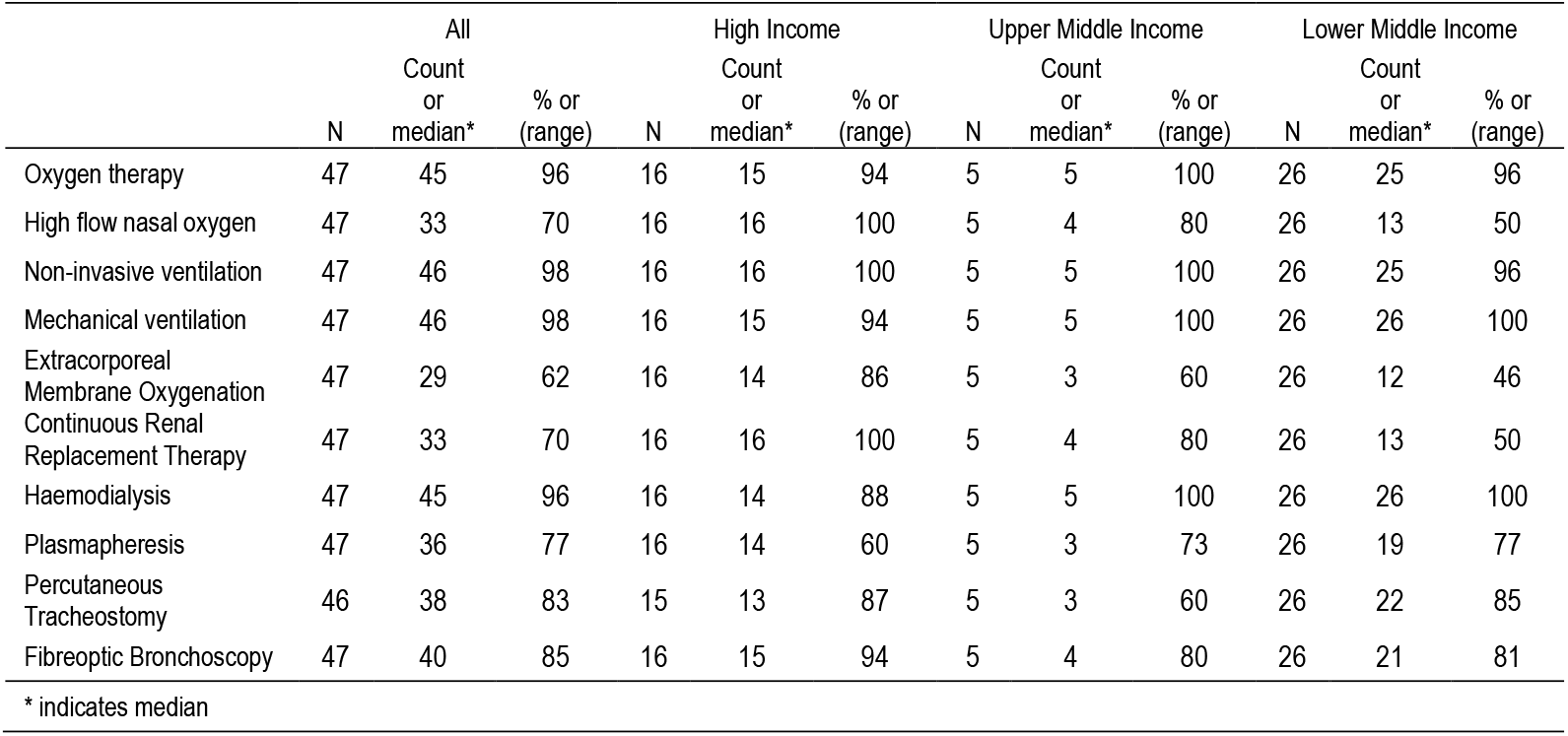
ICU clinical interventions performed per income group

Availability of ICU interventions was similar across income groups except for continuous renal replacement therapy which was only available in 50% LMICs compared to 100% and 80% in HIC and UMIC respectively (Figure 11, Table 5).

**Figure 11.**
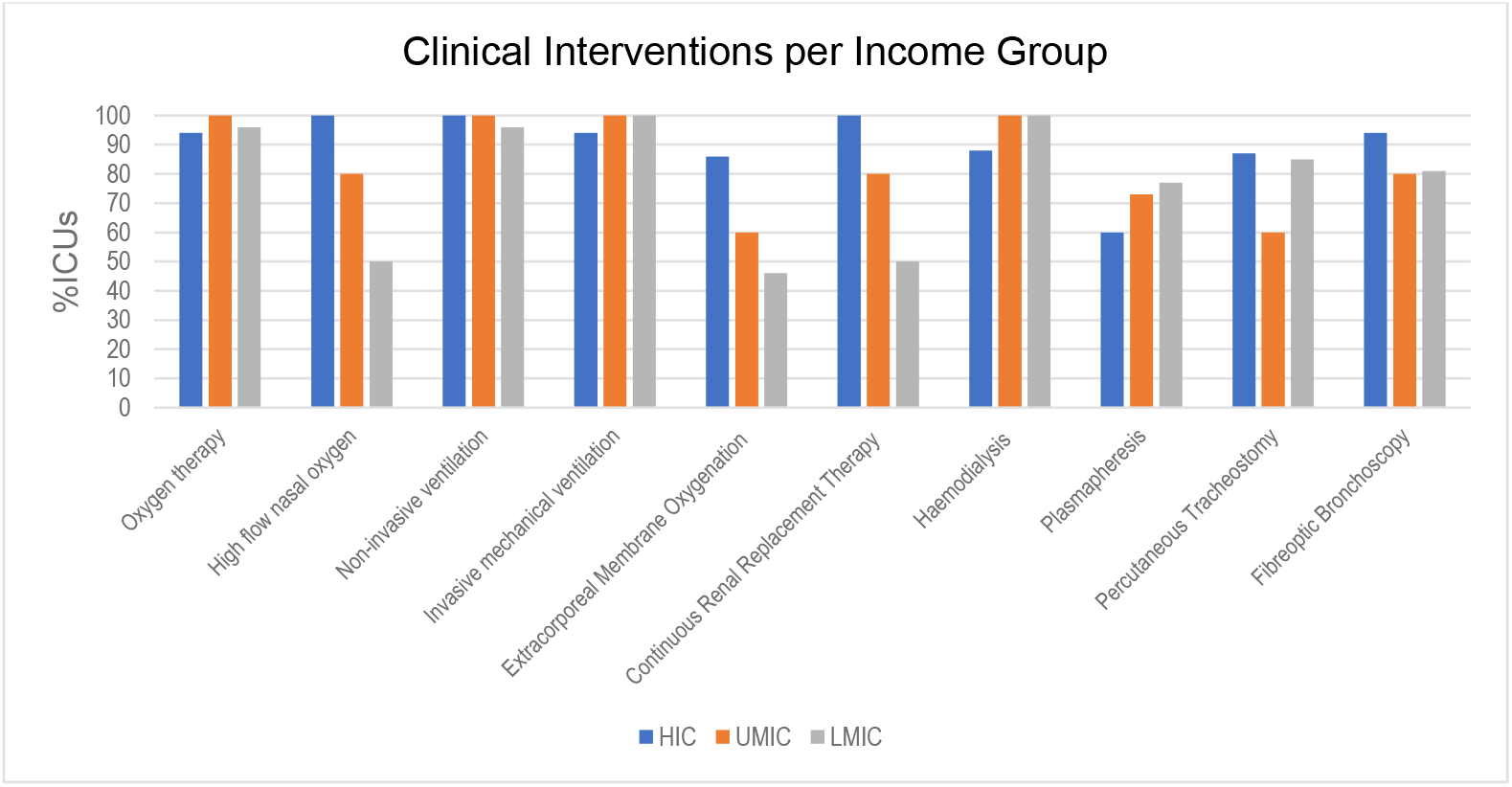
ICU clinical interventions availability according to country income group

Nevertheless, all ICUs had some form of renal replacement therapy available as either continuous renal replacement therapy or haemodialysis. More complex therapies, such as ECMO, were more available in HIC ICUs (86%) compared to UMIC (60%) and LMIC (46%) ICUs.

#### Monitoring

Cardiorespiratory physiological monitoring modalities such as ECG, intra-arterial pressure, central venous pressure (CVP), pulse oximetry (SP0_2_) and end tidal carbon dioxide (E_T_C0_2_) monitoring were standard in around 80% or more of the ICUs across the region (Figure 12, Table 6).

**Figure 12.**
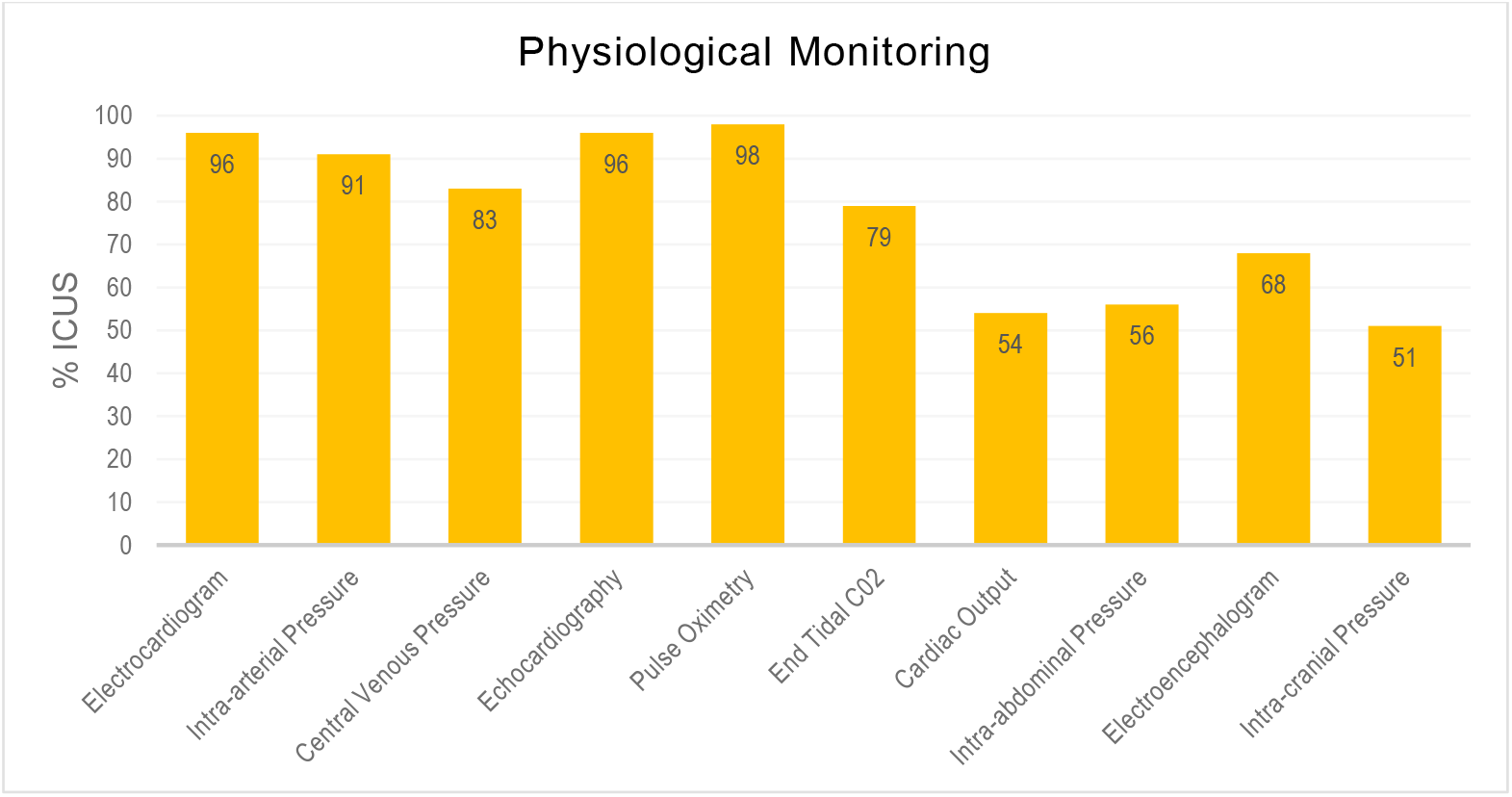
Availability of monitoring modalities in participating ICUs, Asia Pacific region

**Table 6.**
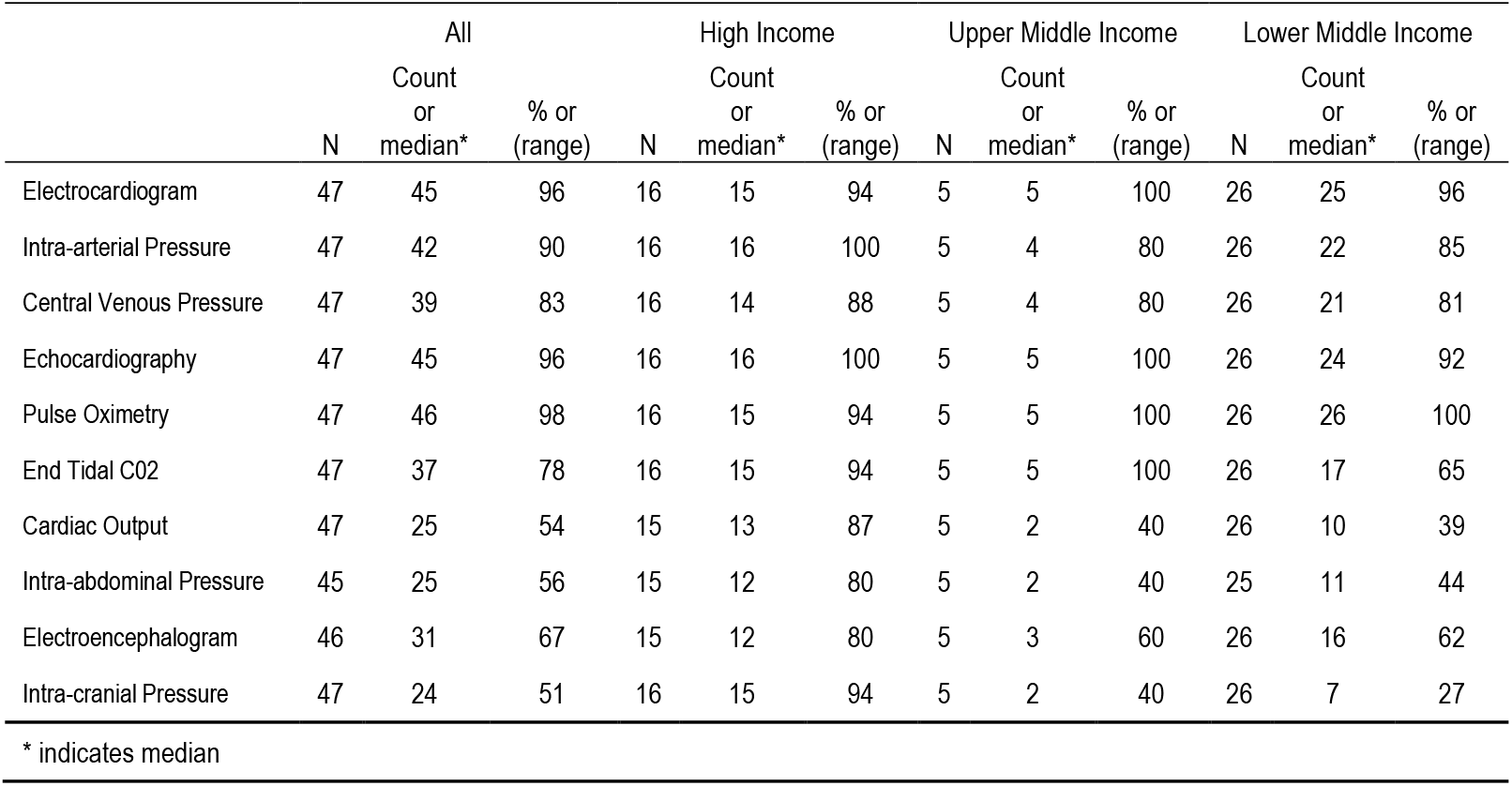
ICU physiological monitoring availability per income group

While more complex modalities such as cardiac output measurement, intra-abdominal pressure and intracranial pressure monitoring were only available within approximately half of the ICUs.

Significant differences were evident between income groups in the use of cardiac output measurement, intraabdominal pressure and intracranial pressure with only 40% or less of ICUs in UMIC and LMICs having access to these modalities (Figure 13, Table 6).

**Figure 13.**
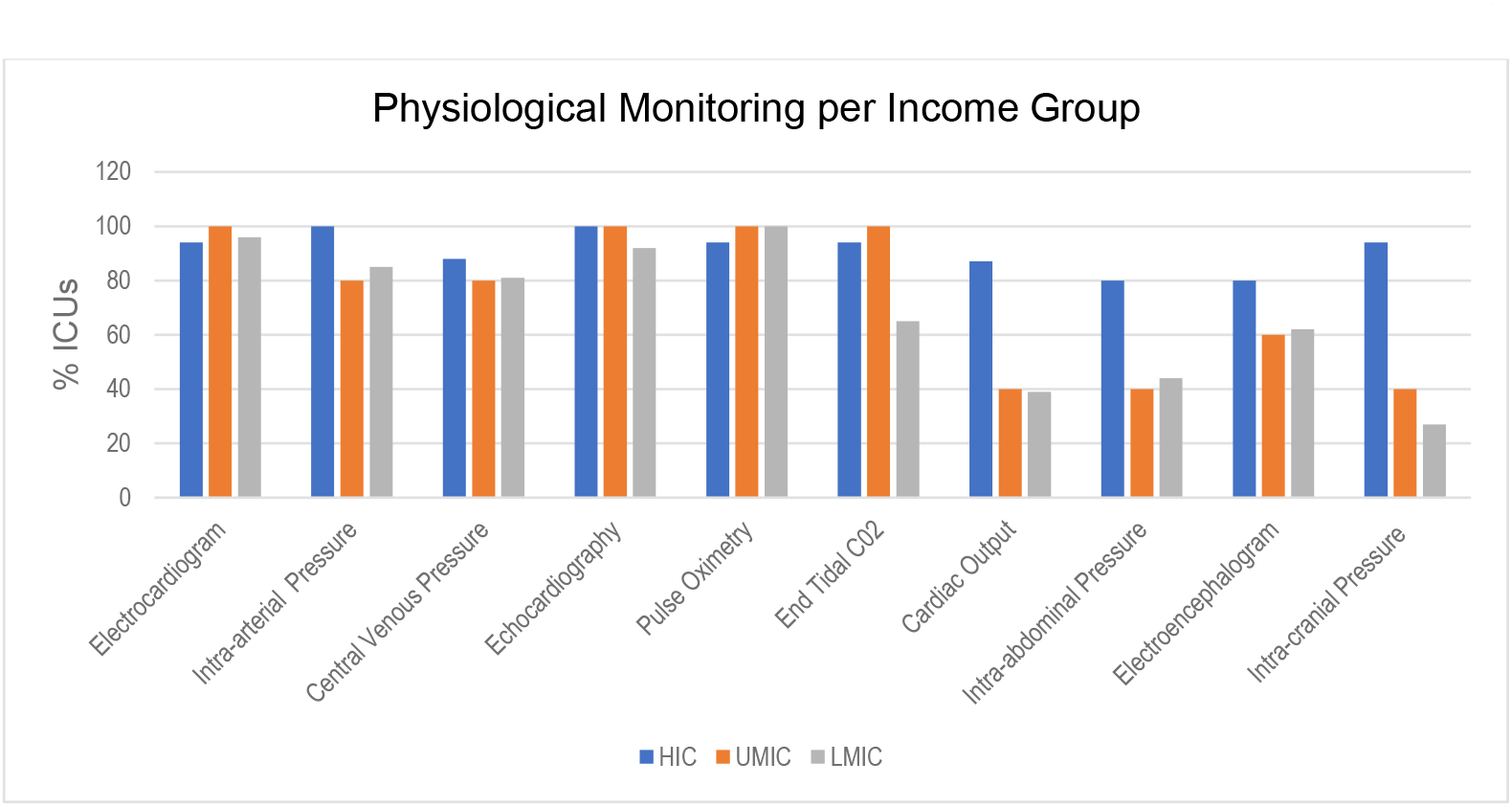
ICU physiological monitoring availability according to country income group

### Pharma therapeutics

#### Intravenous fluids and medications

The availability of intravenous crystalloids, colloids, vasoactive drugs and hydrocortisone in ICU was similar across the region (Figure 14, Table 7).

**Figure 14.**
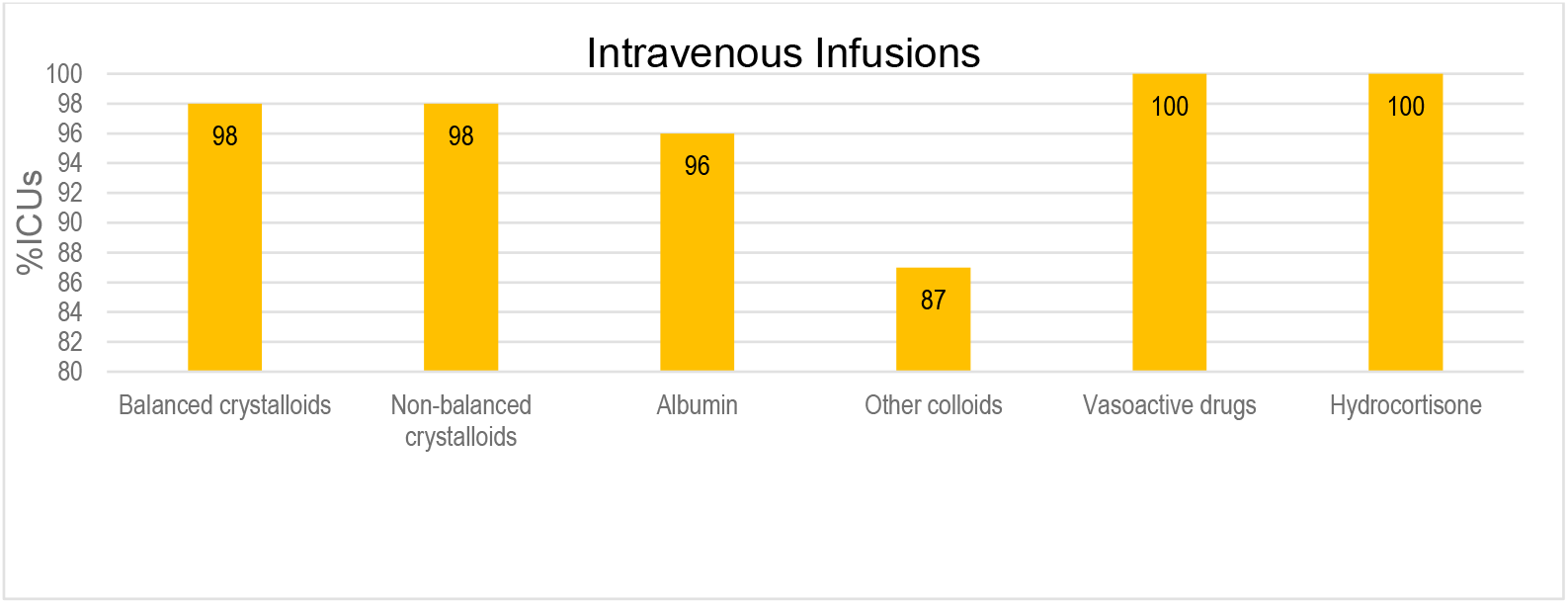
Access to intravenous fluids and medications in participating ICUs, Asia Pacific region

**Table 7.** ICU intravenous fluids and medications availability per income group

|  | All |  |  | High Income |  |  | Upper Middle Income |  |  | Lower Middle Income |  |  |
| --- | --- | --- | --- | --- | --- | --- | --- | --- | --- | --- | --- | --- |
|  | N | Count<br>or<br>median* | % or<br>(range) | N | Count<br>or<br>median* | % or<br>(range) | N | Count<br>or<br>median* | % or<br>(range) | N | Count<br>or<br>median* | % or<br>(range) |
| Balanced crystalloids | 47 | 46 | 98 | 16 | 16 | 100 | 5 | 5 | 100 | 26 | 25 | 96 |
| Non-balanced crystalloids | 47 | 46 | 98 | 16 | 16 | 100 | 5 | 5 | 100 | 26 | 25 | 96 |
| Albumin | 47 | 45 | 96 | 16 | 16 | 100 | 5 | 5 | 100 | 26 | 24 | 92 |
| Other colloids | 47 | 41 | 87 | 16 | 14 | 88 | 5 | 5 | 100 | 26 | 22 | 85 |
| Vasoactive Drugs | 47 | 47 | 100 | 16 | 16 | 100 | 5 | 5 | 100 | 26 | 26 | 100 |
| Hydrocortisone | 47 | 47 | 100 | 16 | 16 | 100 | 5 | 5 | 100 | 26 | 26 | 100 |

\* indicates median

The availability of intravenous crystalloids, colloids, vasoactive drugs and hydrocortisone in ICU was similar across the three income groups with between 85% and 100% of ICUs able to access these resources (Figure 15, Table 7).

**Figure 15.**
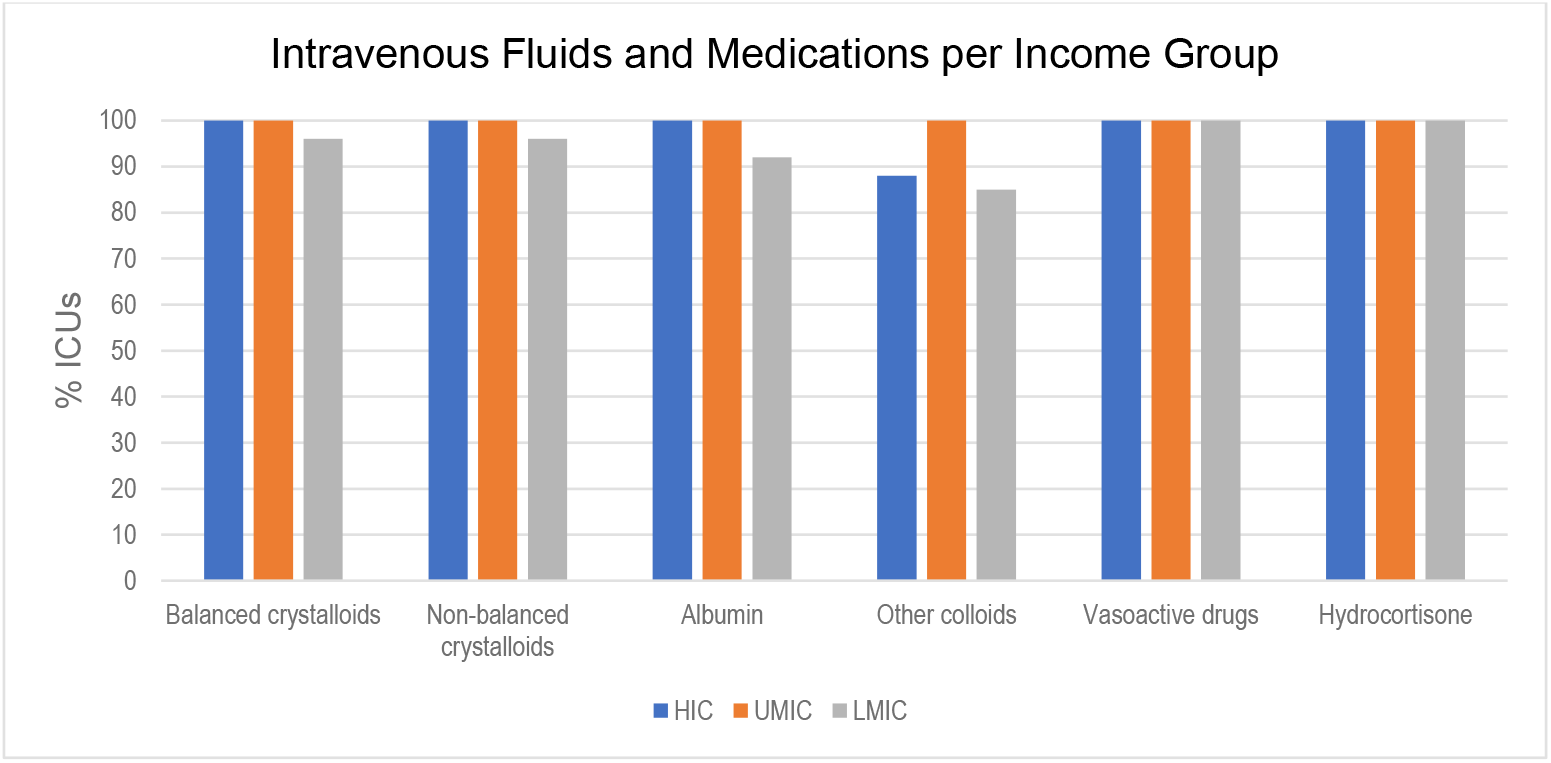
ICU intravenous fluids and medications available according to country income group

#### Antivirals and antimicrobials

The availability of antivirals varied widely depending on the drug in question with only 12% of ICUs reporting access to Remdesevir compared to 88% reporting access to Hydroxychloroquine (Figure 16, Table 8).

**Figure 16.**
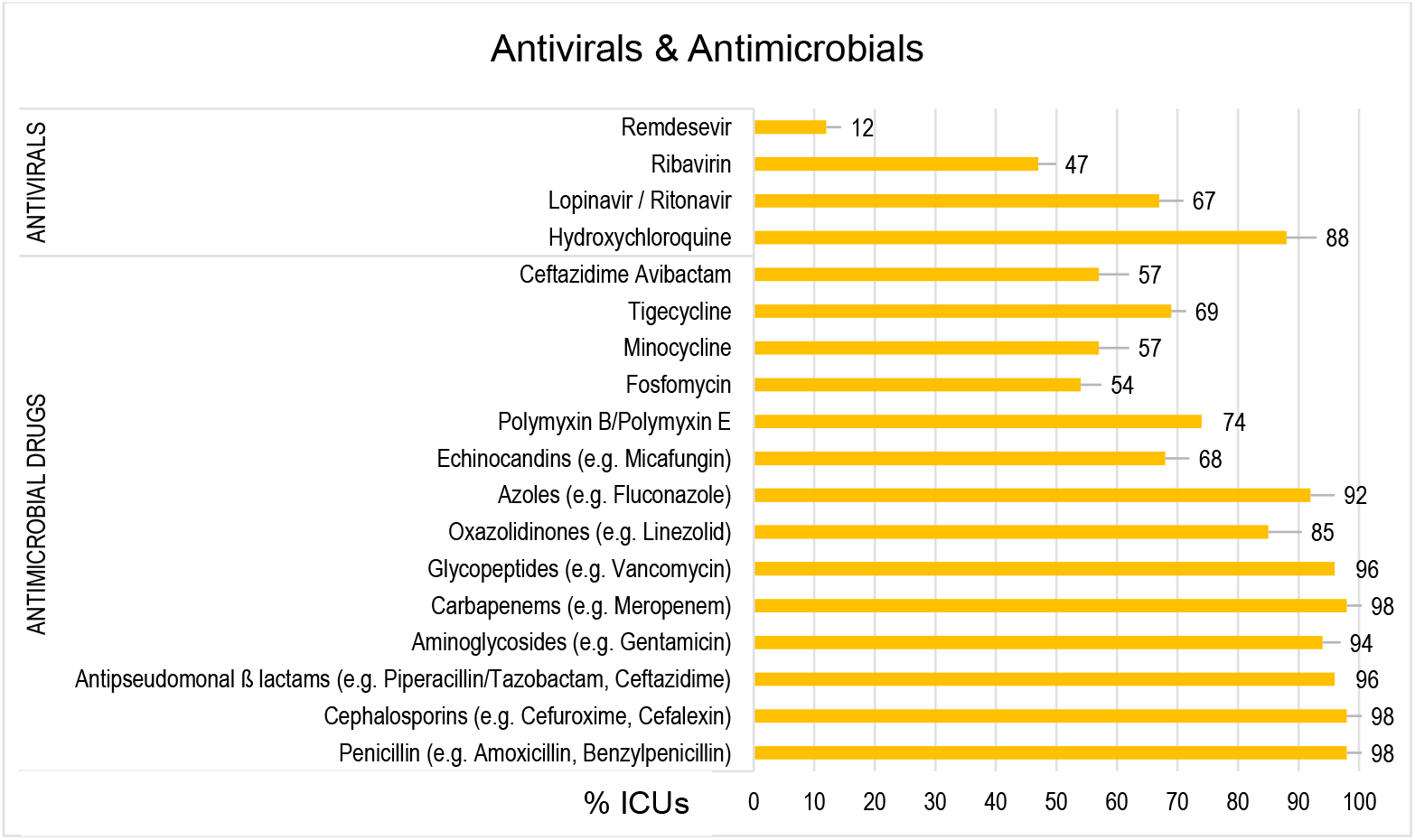
Access to antivirals and antimicrobials in participating ICUs, Asia Pacific region

**Table 8.** Antiviral and antimicrobial availability per income group

|  |  | All |  |  | High Income |  |  | Upper Middle Income |  |  | Lower Middle Income |  |  |
| --- | --- | --- | --- | --- | --- | --- | --- | --- | --- | --- | --- | --- | --- |
|  |  | N | Count<br>or<br>median* | % or<br>(range) | N | Count<br>or<br>median* | % or<br>(range) | N | Count<br>or<br>median* | % or<br>(range) | N | Count<br>or<br>median* | % or<br>(range) |
| Antivirals | Remdesevir | 51 | 6 | 12 | 16 | 5 | 31 | 8 | 0 | 0 | 27 | 1 | 4 |
|  | Ribavirin | 51 | 24 | 47 | 16 | 10 | 63 | 8 | 5 | 63 | 27 | 9 | 33 |
|  | Lopinavir/ritonavir | 51 | 35 | 69 | 16 | 12 | 75 | 8 | 7 | 88 | 27 | 16 | 59 |
|  | Hydroxychloroquine | 51 | 45 | 88 | 16 | 13 | 81 | 8 | 7 | 88 | 27 | 25 | 93 |
| Antimicrobials | Ceftaz-Avibactam | 46 | 26 | 57 | 15 | 11 | 73 | 5 | 1 | 20 | 26 | 14 | 54 |
|  | Tigecycline | 45 | 31 | 69 | 14 | 9 | 64 | 5 | 3 | 60 | 26 | 19 | 73 |
|  | Minocycline | 47 | 27 | 57 | 16 | 13 | 81 | 5 | 1 | 20 | 26 | 13 | 50 |
|  | Fosfomycin | 46 | 25 | 54 | 15 | 7 | 47 | 5 | 3 | 60 | 26 | 15 | 58 |
|  | Polymyxin | 47 | 35 | 75 | 16 | 12 | 75 | 5 | 2 | 40 | 26 | 21 | 81 |
|  | Echinocandins (e.g.<br>Micafungin) | 47 | 32 | 68 | 16 | 14 | 88 | 5 | 3 | 60 | 26 | 15 | 58 |
|  | Azole (e.g. Fluconazole) | 47 | 43 | 92 | 16 | 14 | 88 | 5 | 4 | 80 | 26 | 25 | 96 |
|  | Oxazolidinones (e.g.<br>Linezolid) | 47 | 40 | 85 | 16 | 15 | 94 | 5 | 2 | 40 | 26 | 23 | 89 |
|  | Glycopeptides (e.g.<br>Vancomycin) | 49 | 45 | 96 | 16 | 15 | 94 | 5 | 4 | 80 | 26 | 26 | 100 |
|  | Carbapenems (e.g.<br>Meropenem) | 47 | 46 | 98 | 16 | 16 | 100 | 5 | 4 | 80 | 26 | 26 | 100 |
|  | Aminoglycoside (e.g.<br>Gentamicin) | 47 | 44 | 94 | 16 | 15 | 94 | 5 | 4 | 80 | 26 | 25 | 96 |
|  | Anti-pseudo Beta lactam | 47 | 45 | 96 | 16 | 15 | 94 | 5 | 5 | 100 | 26 | 25 | 96 |
|  | Cephalosporin (e.g.<br>Cefuroxime, Cefalexin) | 47 | 46 | 98 | 16 | 15 | 94 | 5 | 5 | 100 | 26 | 26 | 100 |
|  | Penicillin | 47 | 46 | 98 | 16 | 15 | 94 | 5 | 5 | 100 | 26 | 26 | 100 |

\* indicates median

A wide variety of antimicrobials, listed as essential medicine by the World Health Organisation (WHO) such as gentamicin and amoxicillin and cefuroxime, were available in most units across the region. However, “reserve” antimicrobials classified in the WHO Access, Watch, and Reserve (AWaRE) tool^6^, such as Ceftazidime Avibactam, Minocycline and Fosfomycin, were limited.

Antiviral availability across income groups was similar though UMIC and LMIC ICUs had limited to no access to Remdesevir, 0% and 4% respectively, with LMIC having the best access to Hydro chloroquine (93%) (Figure 17, Table 8).

**Figure 17.**
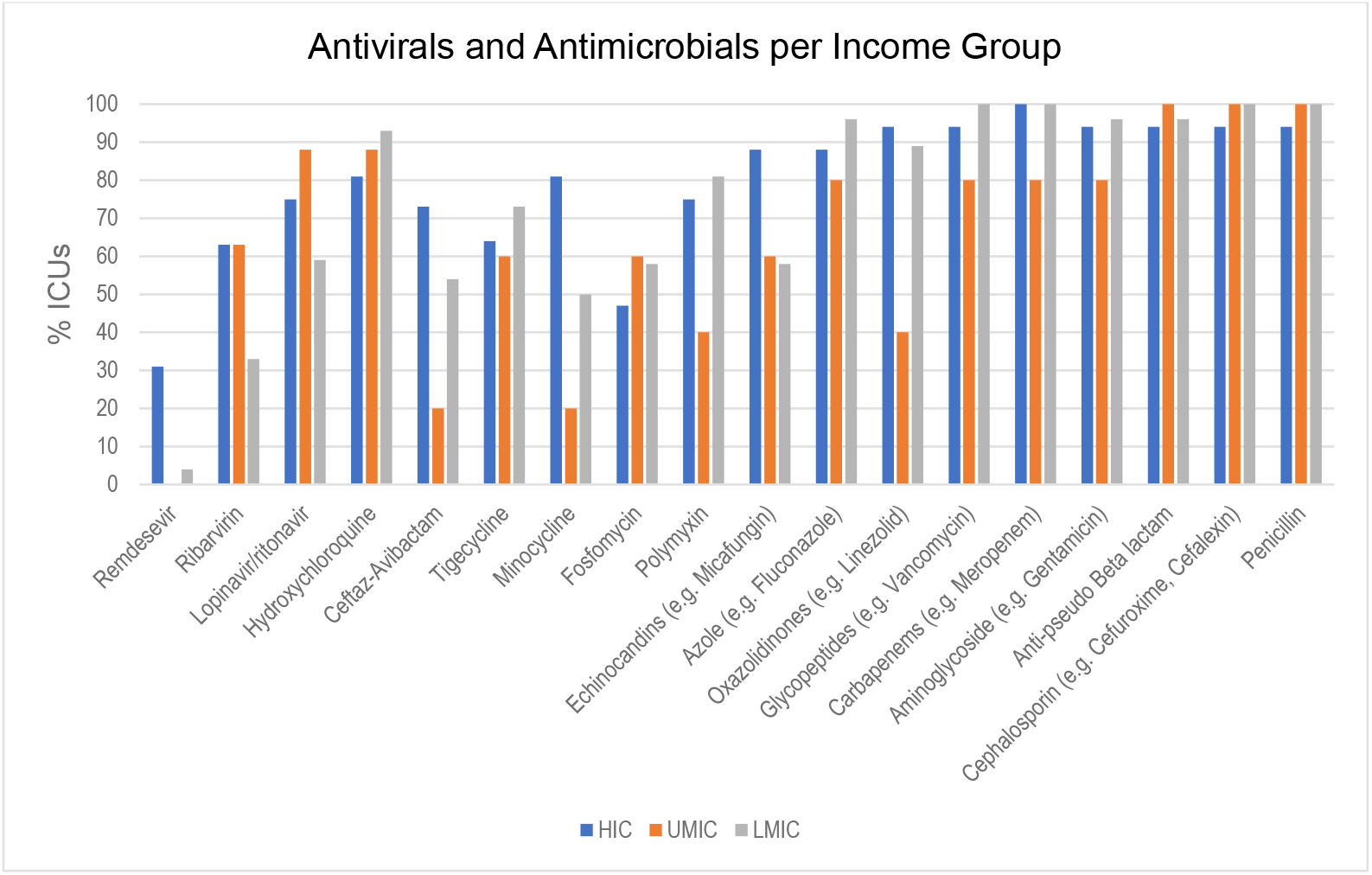
Antiviral and antimicrobial availability according to country income group

LMICs in general had less availability of antimicrobials than HIC but ICU’s in UMIC recorded the lowest access to a number of antimicrobials including Ceftaz-Avibactam (20%), Tigecycline (60%), Minocycline (20%), Polymyxin (40%) and Linezolid (40%).

### Sepsis management

#### Evidence based guidelines

Sepsis specific management guidelines were used broadly across the region, with 86% of ICUs reporting the availability of international, national, state/region and unit specific guidelines (Figure 18, Table 18). Despite the large proportion of LMIC in the survey only 14% of ICUs overall used the tailored LMIC sepsis management guidelines. Clinicians reported sufficient resources and training to adhere to guidelines in 78% of ICU’s.

**Figure 18.**
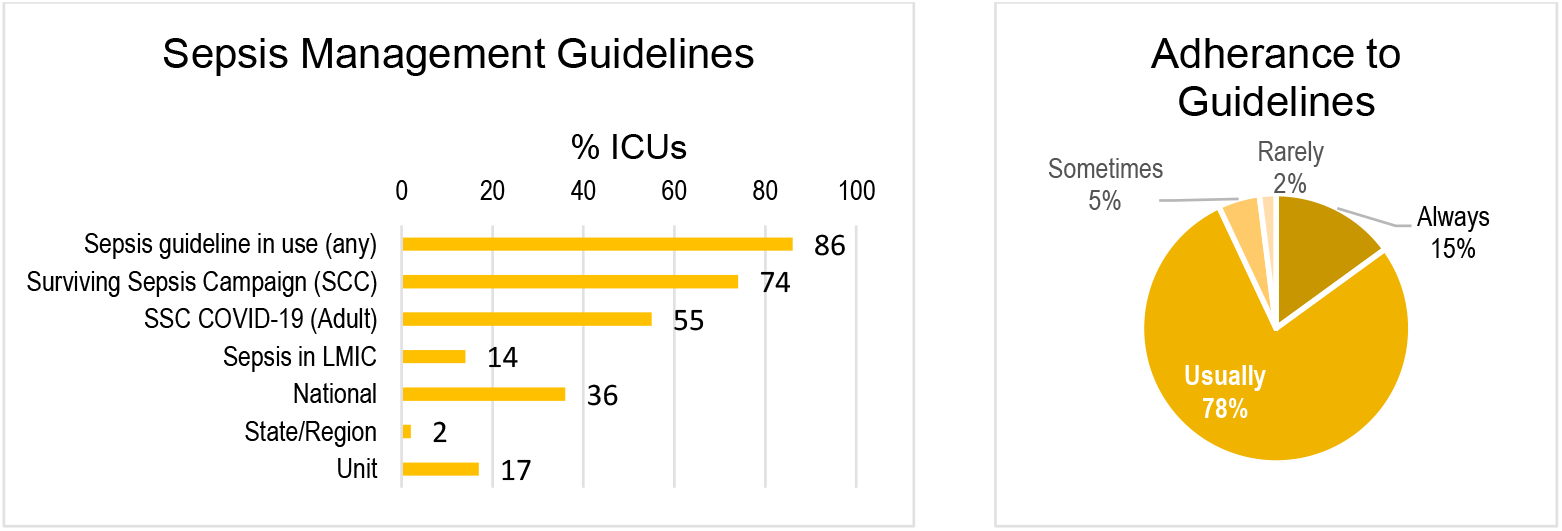
Sepsis guidelines used and adherence in participating ICUs, Asia Pacific region

More UMIC (100%) and LMIC ICU’s (92%) had sepsis guidelines in place than HIC ICU’s (67%). The Surviving Sepsis Campaign (SSC) guidelines were most commonly used across all income groups (Figure 19, Table 9).

**Figure 19.**
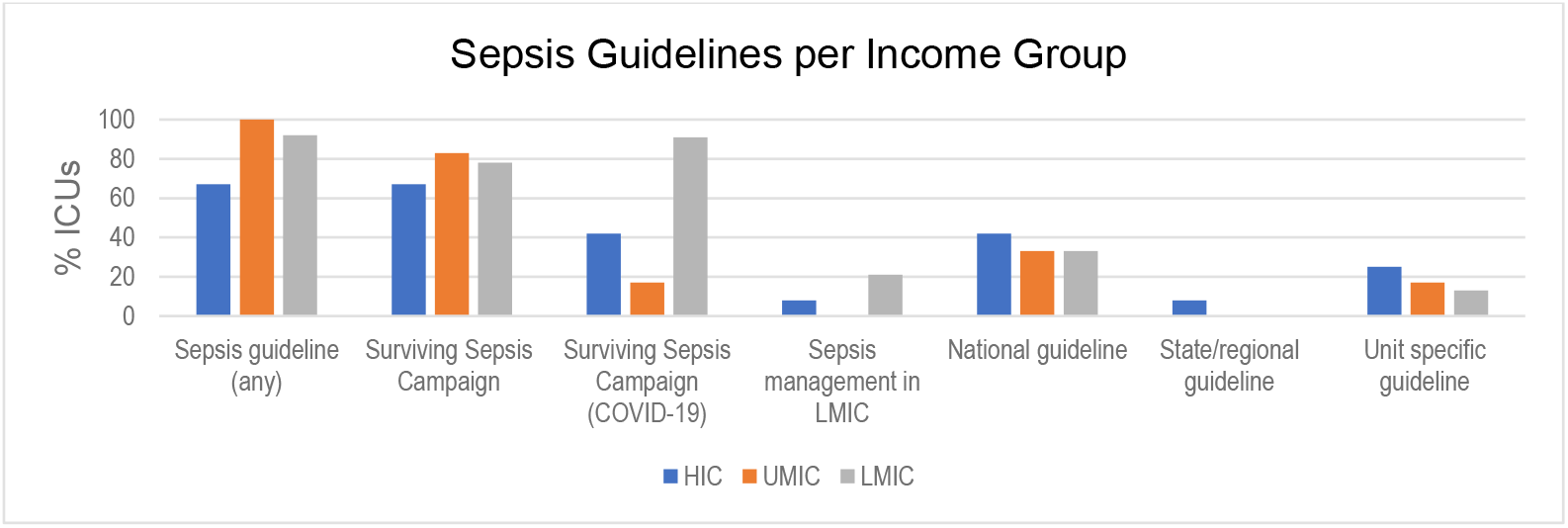
Sepsis guidelines in use according to country income group

**Table 9.**
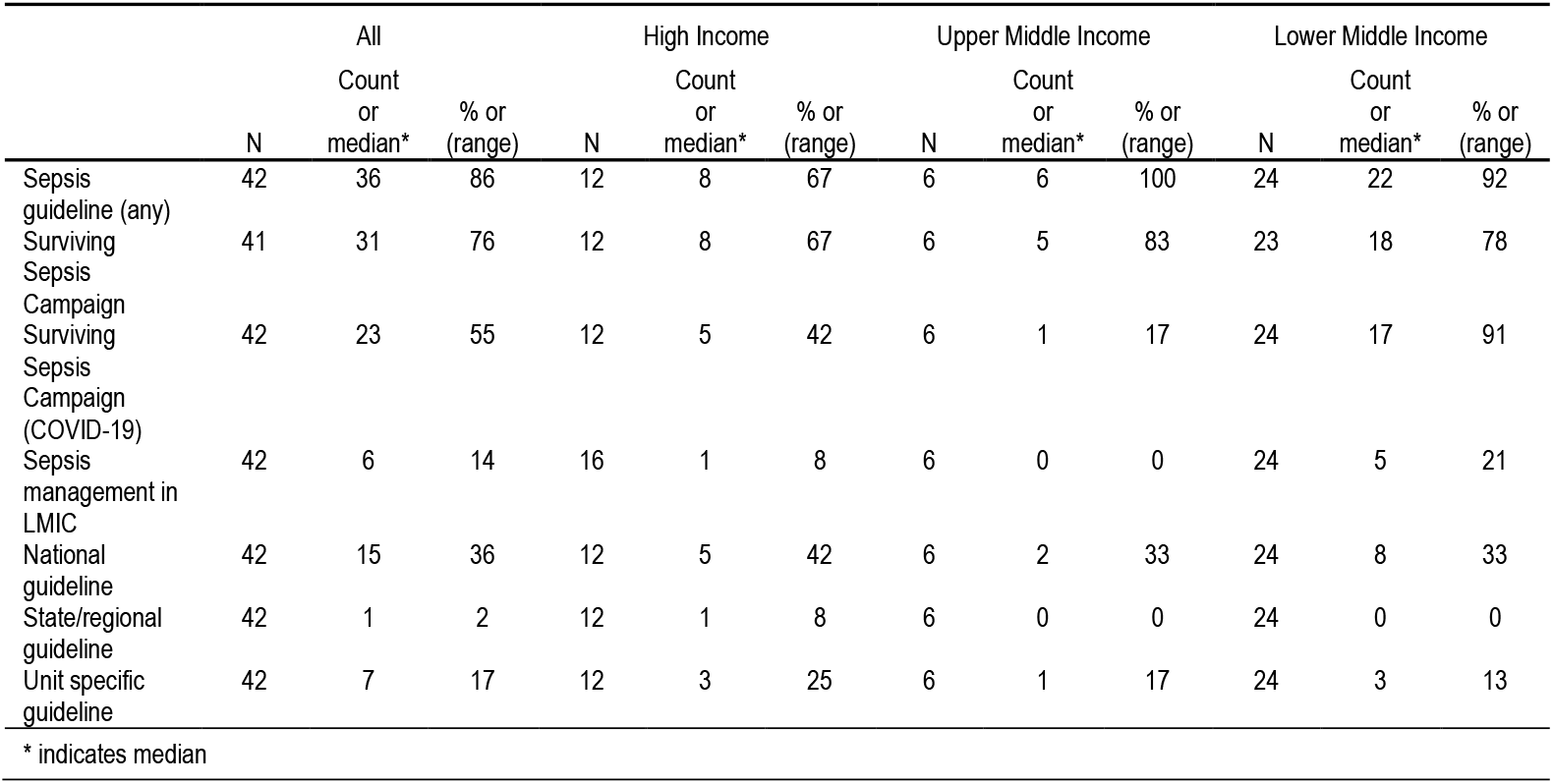
Sepsis Guidelines per Income Group

|  | All |  |  | High Income |  |  | Upper Middle Income |  |  | Lower Middle Income |  |  |
| --- | --- | --- | --- | --- | --- | --- | --- | --- | --- | --- | --- | --- |
|  | N | Count or median* | % or (range) | N | Count or median* | % or (range) | N | Count or median* | % or (range) | N | Count or median* | % or (range) |
| Sepsis guideline (any) | 42 | 36 | 86 | 12 | 8 | 67 | 6 | 6 | 100 | 24 | 22 | 92 |
| Surviving Sepsis Campaign | 41 | 31 | 76 | 12 | 8 | 67 | 6 | 5 | 83 | 23 | 18 | 78 |
| Surviving Sepsis Campaign (COVID-19) | 42 | 23 | 55 | 12 | 5 | 42 | 6 | 1 | 17 | 24 | 17 | 91 |
| Sepsis management in LMIC | 42 | 6 | 14 | 16 | 1 | 8 | 6 | 0 | 0 | 24 | 5 | 21 |
| National guideline | 42 | 15 | 36 | 12 | 5 | 42 | 6 | 2 | 33 | 24 | 8 | 33 |
| State/regional guideline | 42 | 1 | 2 | 12 | 1 | 8 | 6 | 0 | 0 | 24 | 0 | 0 |
| Unit specific guideline | 42 | 7 | 17 | 12 | 3 | 25 | 6 | 1 | 17 | 24 | 3 | 13 |
| * indicates median |  |  |  |  |  |  |  |  |  |  |  |  |

There was relatively poor uptake of the SCC COVID-19 guidelines in HIC (55%) and UMIC (17%) ICUs compared to LMIC ICUs (71%). In contrast to expectations, only 21% of LMIC ICU’s were using the adapted SSC guidelines for LMIC’s, which were not used UMIC and only 14% of HIC ICU’s. Around 30-40% of all income groups used a national guideline for sepsis management.

### Quality and research

Infection control policies and antimicrobial stewardship programs (AMS) were available in 98% and 88% of units respectively (Figure 20, Table 10). However, only 56% of responding ICUs reported the availability of a specific sepsis consultation service available. Unit based and multicentre research was conducted in 65% and 52% respectively.

**Figure 20.**
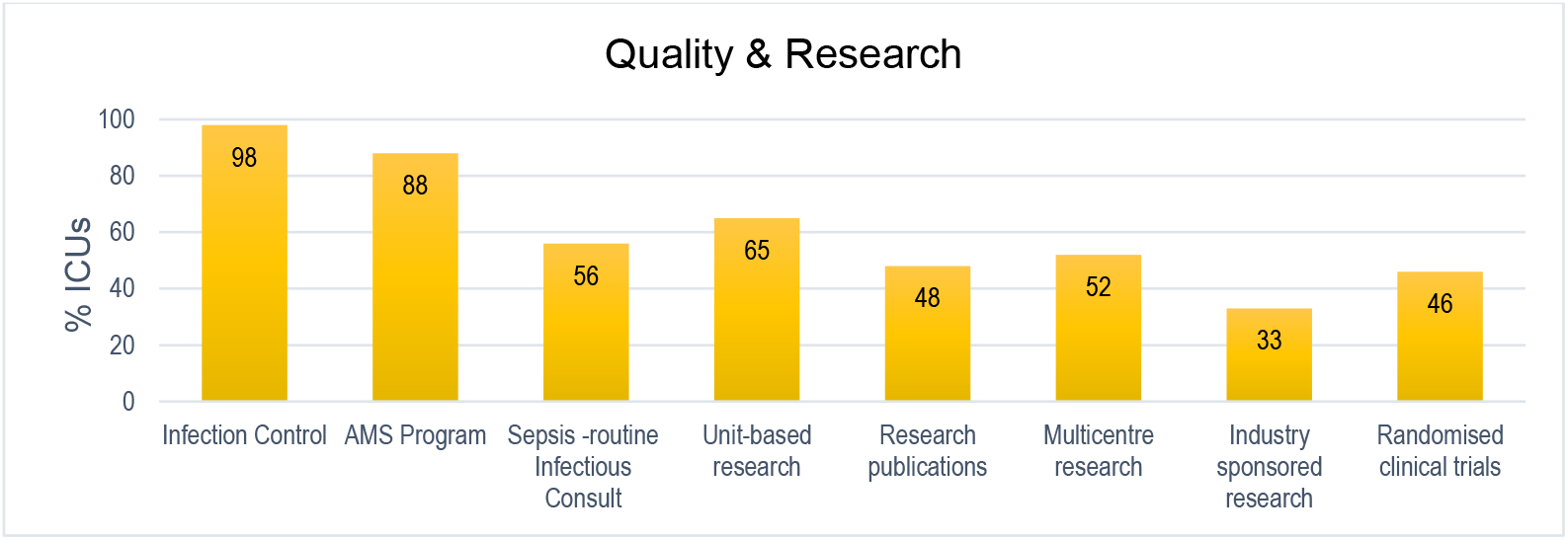
Quality improvement and research in participating ICUs, Asia Pacific region

**Table 10.** Quality improvement activities and clinical research per income group

|  | All |  |  | High Income |  |  | Upper Middle Income |  |  | Lower Middle Income |  |  |
| --- | --- | --- | --- | --- | --- | --- | --- | --- | --- | --- | --- | --- |
|  | N | Count or median* | % or (range) | N | Count or median* | % or (range) | N | Count or median* | % or (range) | N | Count or median* | % or (range) |
| Infection control policies | 48 | 47 | 98 | 16 | 16 | 100 | 6 | 6 | 100 | 26 | 25 | 96 |
| AMS program | 48 | 42 | 88 | 16 | 14 | 88 | 6 | 6 | 100 | 26 | 22 | 85 |
| Sepsis consult (routine) | 58 | 27 | 56 | 16 | 10 | 63 | 6 | 1 | 17 | 26 | 16 | 62 |
| Unit-based research | 58 | 31 | 65 | 16 | 11 | 69 | 6 | 4 | 67 | 26 | 16 | 62 |
| Research publications | 58 | 23 | 48 | 16 | 11 | 69 | 4 | 2 | 33 | 26 | 10 | 39 |
| Multicentre research | 48 | 25 | 52 | 16 | 10 | 63 | 6 | 1 | 17 | 26 | 14 | 54 |
| Industry research | 48 | 16 | 33 | 16 | 7 | 44 | 6 | 1 | 17 | 26 | 8 | 31 |
| RCT's | 48 | 22 | 46 | 16 | 9 | 56 | 6 | 2 | 33 | 26 | 11 | 42 |
| * indicates median |  |  |  |  |  |  |  |  |  |  |  |  |

Infection control policies and AMS programs operated in ICUs across all income groups (Figure 21, Table 10). Routine infectious team consult for sepsis patients was not as ubiquitous with only 17% of UMIC ICU’s having adopted this quality strategy compared to HIC (63%) and LMIC (62%) ICU’s. A similar proportion of ICU’s conducted unit-based research across all income groups (range 62%-69%) but UMIC reported considerably less multicentre research (17%), industry sponsored research (17%) and RCT’s (33%).

**Figure 21.**
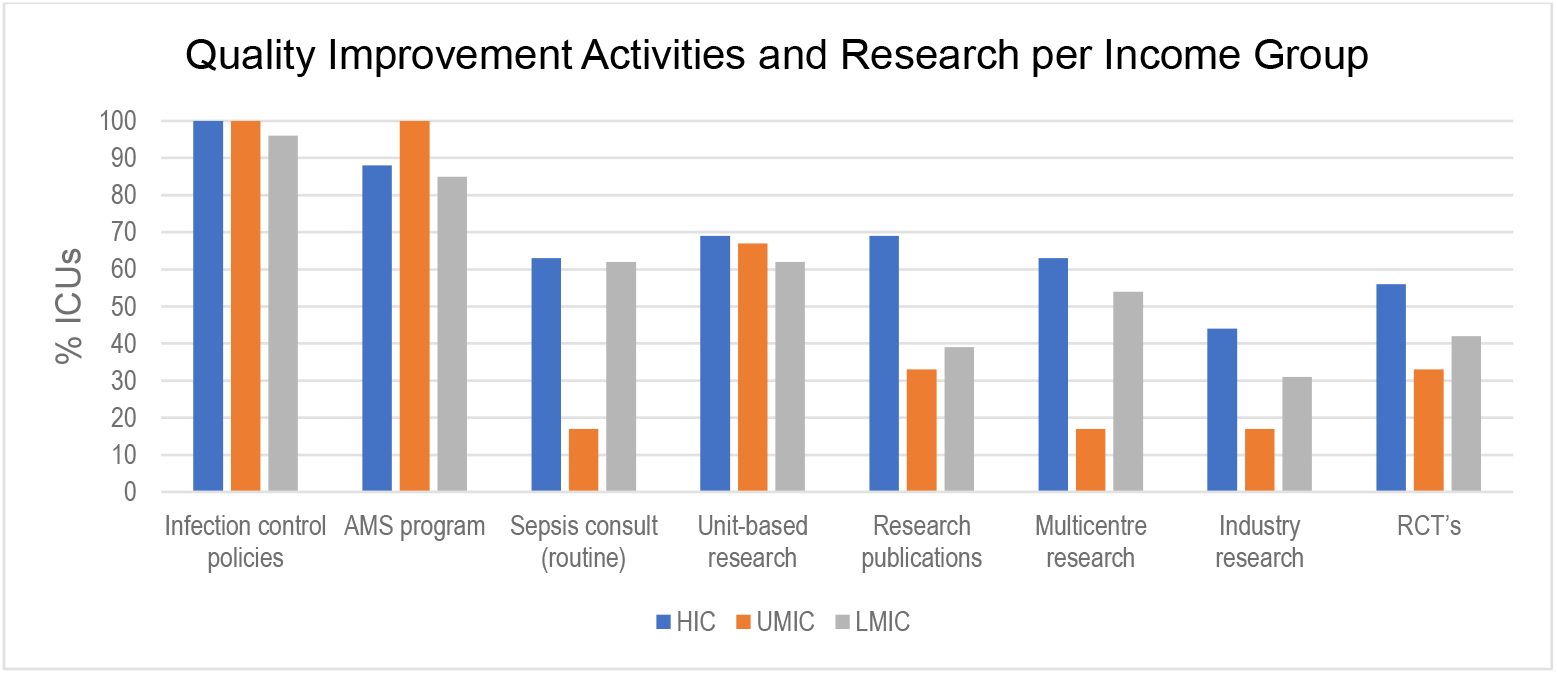
Quality improvement and research according to country income group

### SARS-CoV2 and disaster preparedness

#### Resources and planning

All units reported a good availability of basic equipment (Personal protection equipment, N95 masks, gloves, alcohol gel) (Figure 22, Table 11).

**Figure 22.**
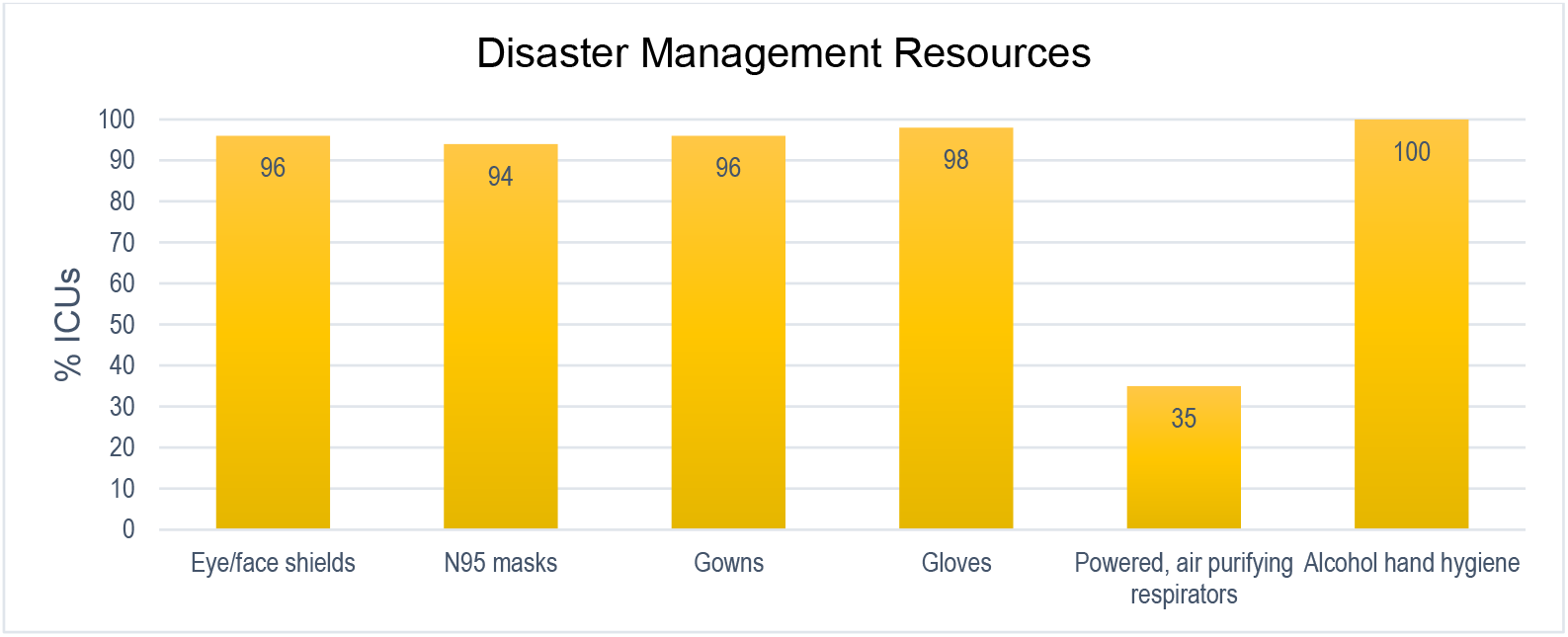
Resourcing for disaster management in participating ICUs, Asia Pacific region

**Table 11.** Disaster Management Resources per Income Group

|  | All |  |  | High Income |  |  | Upper Middle Income |  |  | Lower Middle Income |  |  |
| --- | --- | --- | --- | --- | --- | --- | --- | --- | --- | --- | --- | --- |
|  | N | Count or median* | % or (range) | N | Count or median* | % or (range) | N | Count or median* | % or (range) | N | Count or median* | % or (range) |
| Eye/face shields | 59 | 56 | 95 | 18 | 16 | 89 | 8 | 8 | 100 | 33 | 32 | 97 |
| N95 | 51 | 48 | 94 | 16 | 16 | 100 | 7 | 7 | 100 | 28 | 25 | 89 |
| Gown | 51 | 49 | 96 | 16 | 15 | 94 | 7 | 7 | 100 | 28 | 27 | 96 |
| Gloves | 51 | 50 | 98 | 16 | 16 | 100 | 7 | 7 | 100 | 28 | 27 | 96 |
| Respirator/purifier | 51 | 18 | 35 | 16 | 7 | 44 | 7 | 4 | 57 | 28 | 7 | 25 |
| Alcohol hand hygiene | 51 | 51 | 100 | 16 | 16 | 100 | 7 | 7 | 100 | 28 | 28 | 100 |

\* indicates median

Personal protection equipment (PPE) for disaster management was readily accessible across all income groups (Figure 23, Table 11) other than air purifying equipment which was available in only half of HIC and UMIC ICU’s and only 25% of LMIC ICU’s.

**Figure 23.**
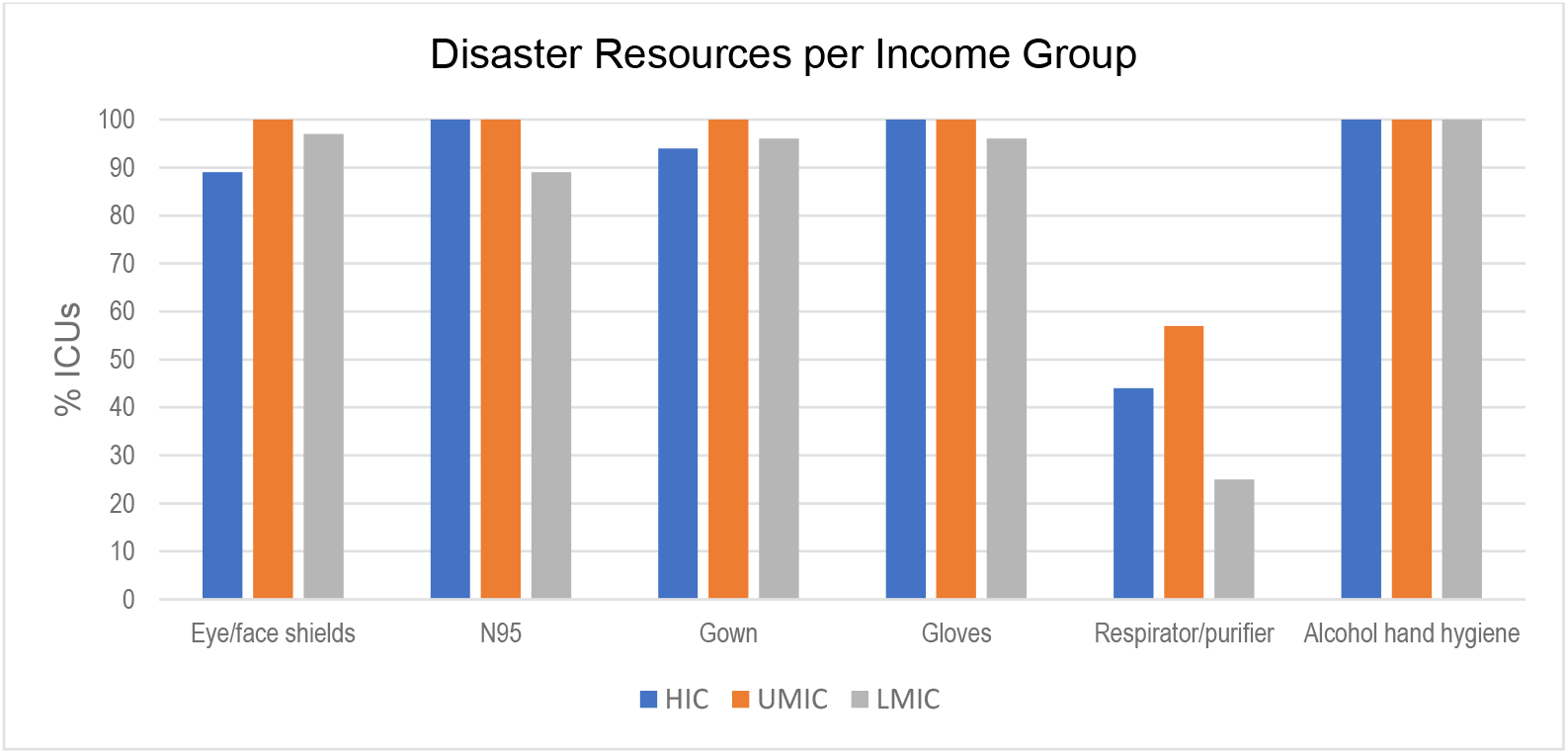
Disaster management resources according to country income group

A majority of respondents had a disaster plan (98%) that included guidelines or training for use of PPE (95%)and patient triage screening and isolation (93%) (Figure 24, Table 12). Bed surge and workforce management were both included in 97% of plans but only 71% included triage surge guidelines and 75% involved simulation training.

**Figure 24.**
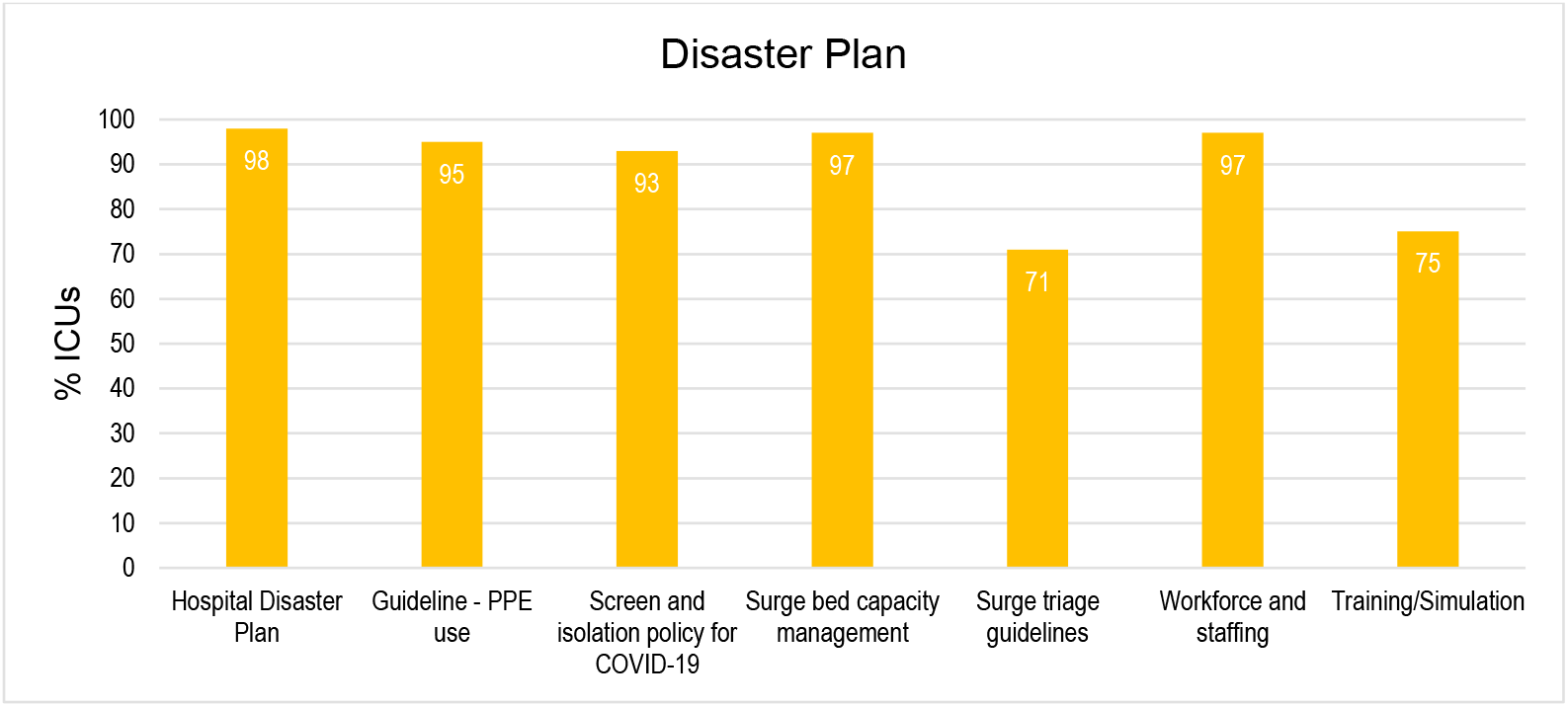
Disaster plan components across participating ICUs, Asia Pacific region

**Table 12.**
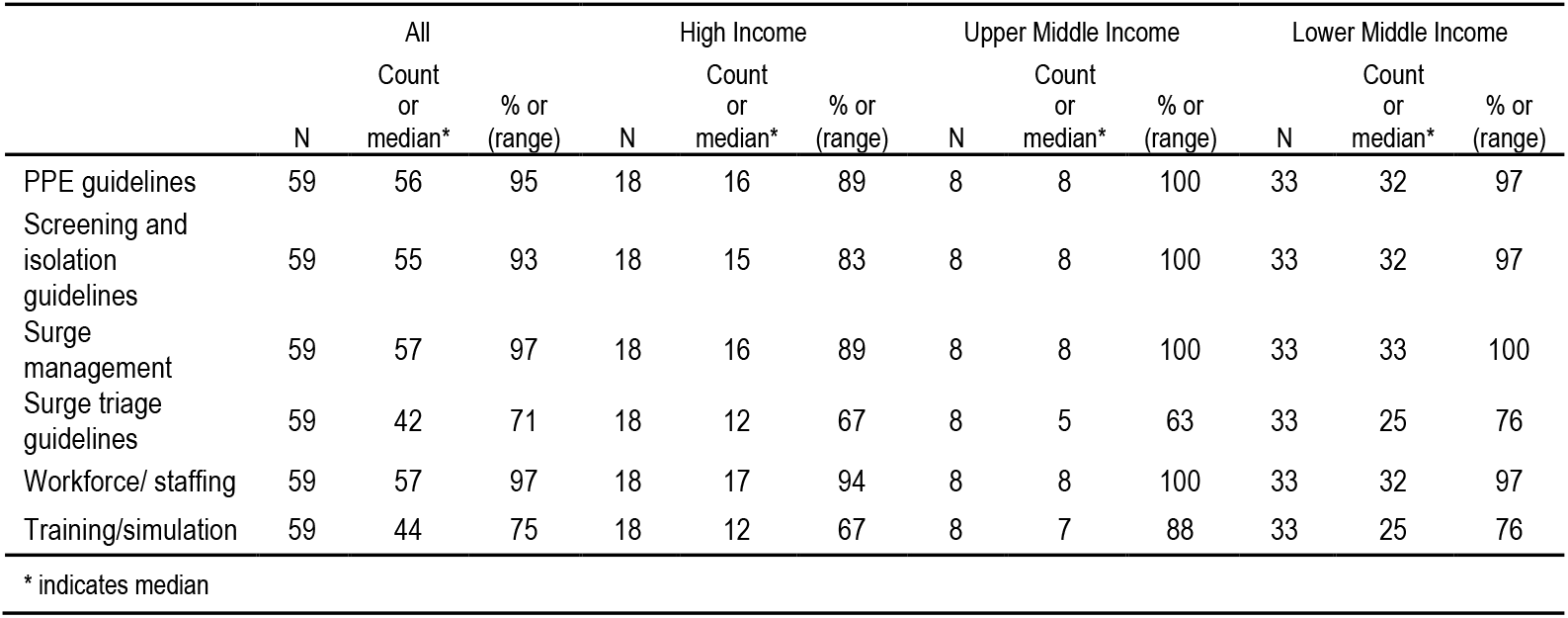
Disaster planning components and preparation per income group

|  | All |  |  | High Income |  |  | Upper Middle Income |  |  | Lower Middle Income |  |  |
| --- | --- | --- | --- | --- | --- | --- | --- | --- | --- | --- | --- | --- |
|  | N | Count or median* | % or (range) | N | Count or median* | % or (range) | N | Count or median* | % or (range) | N | Count or median* | % or (range) |
| PPE guidelines | 59 | 56 | 95 | 18 | 16 | 89 | 8 | 8 | 100 | 33 | 32 | 97 |
| Screening and isolation guidelines | 59 | 55 | 93 | 18 | 15 | 83 | 8 | 8 | 100 | 33 | 32 | 97 |
| Surge management | 59 | 57 | 97 | 18 | 16 | 89 | 8 | 8 | 100 | 33 | 33 | 100 |
| Surge triage guidelines | 59 | 42 | 71 | 18 | 12 | 67 | 8 | 5 | 63 | 33 | 25 | 76 |
| Workforce/ staffing | 59 | 57 | 97 | 18 | 17 | 94 | 8 | 8 | 100 | 33 | 32 | 97 |
| Training/simulation | 59 | 44 | 75 | 18 | 12 | 67 | 8 | 7 | 88 | 33 | 25 | 76 |

\* indicates median

Hospital disaster plans addressed PPE use, screening and isolation, surge management and training across all income groups (Figure 25, Table 12). Fewer plans included surge triage guidelines in HIC (67%) and UMIC (63%) hospitals than in LMIC’s (76%), and considerably less HIC hospitals conducted training (67%) than UMIC (88%) and LMIC (76%) hospitals.

**Figure 25.**
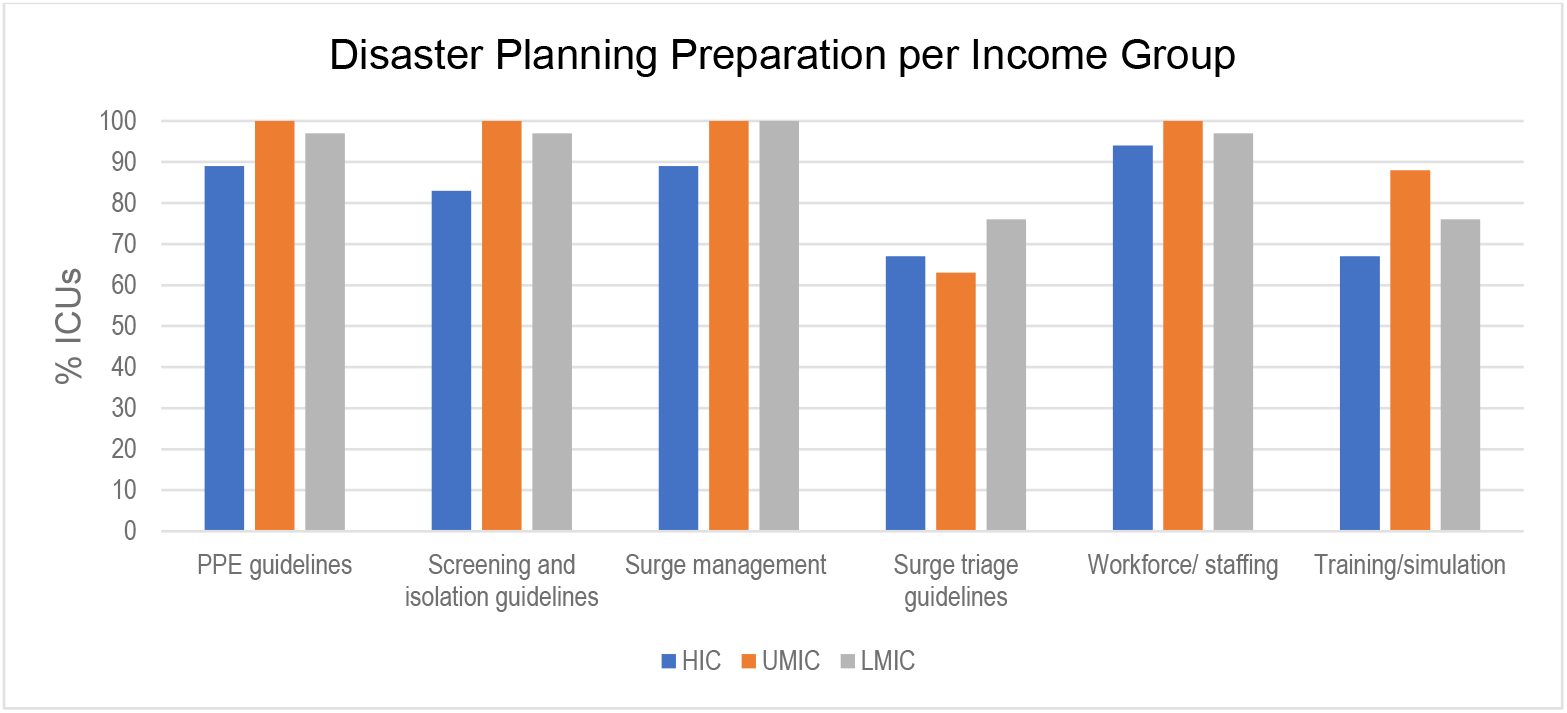
Disaster planning components and preparation according to country income group

#### Isolation and surge capacity

Across the region there was a median of nine single hospital rooms (range 1-25) and seven single ICU rooms (range 1-25) reported (Figure 26, Table 13).

**Figure 26.**
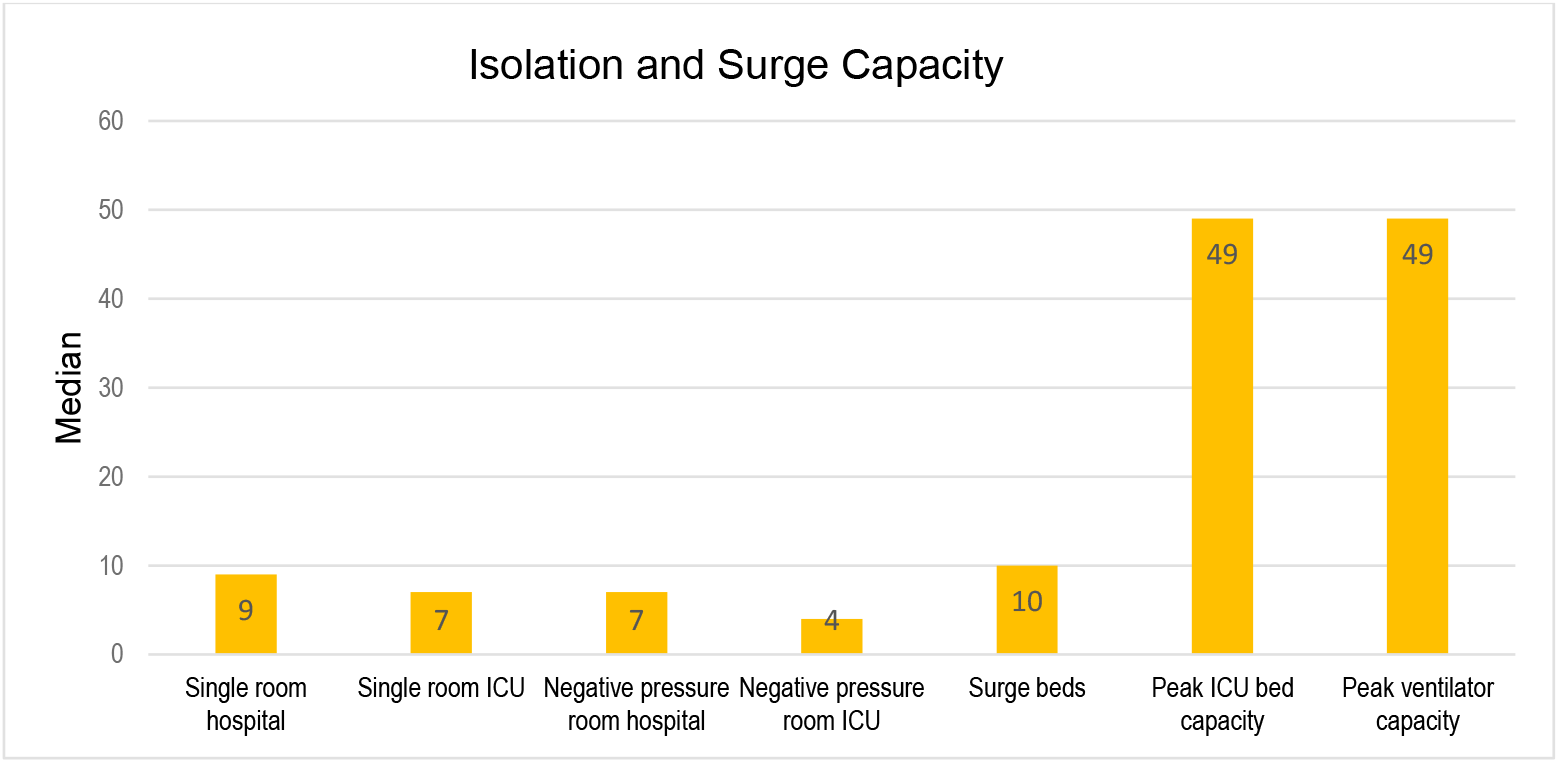
Isolation and surge capacity in participating hospitals, Asia Pacific region

**Table 13.**
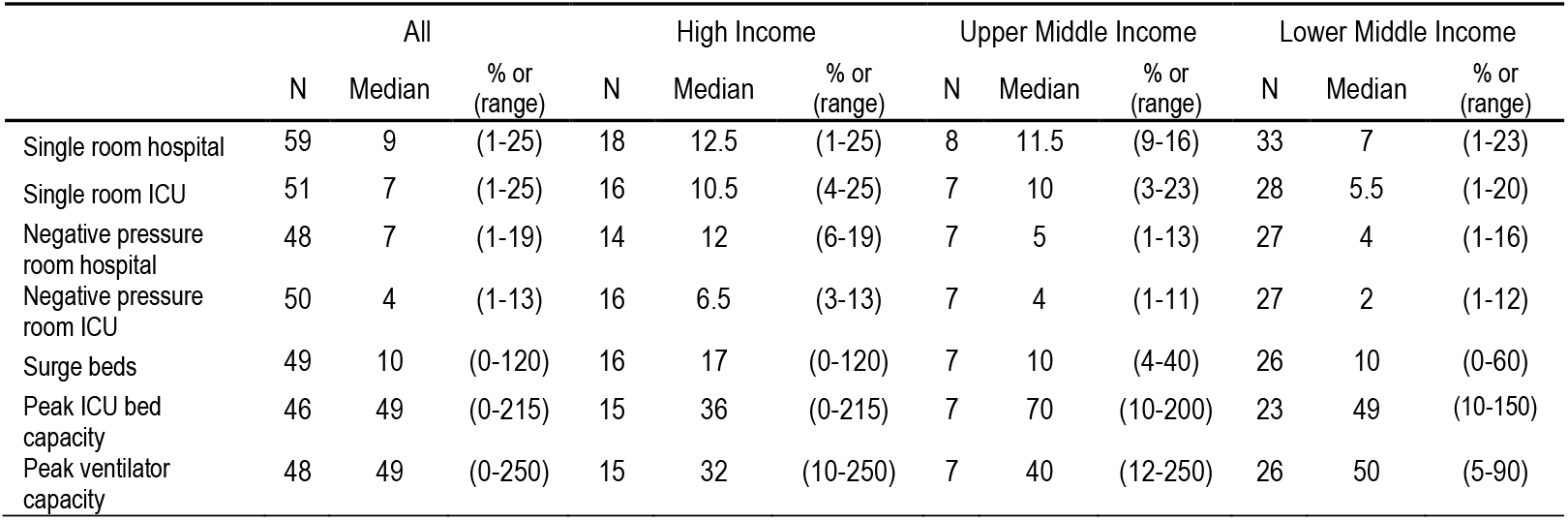
Isolation and surge capacity per income group

The LMIC group reported considerably less hospital and ICU isolation capacity, median 7 beds and 5.5 beds respectively (Figure 27, Table 13). This was also evident for hospital and ICU negative pressure rooms with a median of 4 rooms and 2 rooms respectively in LMICs, approximately half that of UMIC and one quarter or less capacity than in HIC’s. Across the region there was a median surge capacity of 10 ICU beds (range 0-120) reported though the median for HIC’s was 17 surge ICU beds. Peak ventilation capacity was relatively similar for all income groups with a median of 49 (range 0-250) mechanical ventilators.

**Figure 27.**
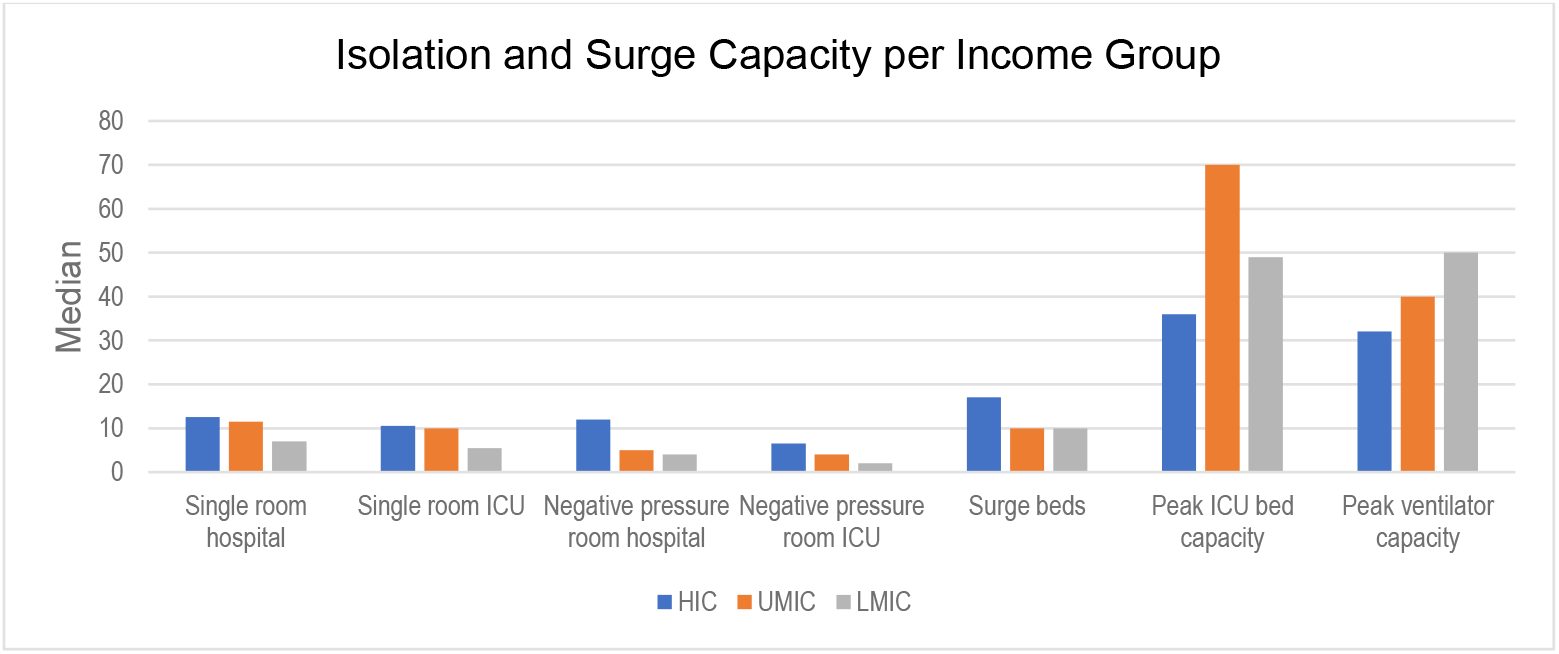
Isolation and surge capacity according to country income group

### Conclusion

Although this survey mostly sampled tertiary ICUs across HICs and LMICs, provision of critical care for management of sepsis varied across the Asia Pacific region. Specific differences include nurse to patient ratios and availability of allied health services.

Conventional organ support modalities such as mechanical ventilation and non-invasive ventilation were commonly available. Even advanced life support like ECMO was available in at least 60% of surveyed ICUs. However, in contrast, essential monitoring devices like EtCO2 were not universally available.

In terms of pandemic and disaster planning, whilst basic personal protective equipment was widely available, LMICs had much lower provisions for isolation and surge capacity. Most ICUs use the SSC guidelines or the adapted SSC guidelines for LMICs, though only 21% of LMIC ICU’s used the adapted version of the SSC guidelines. Essential antimicrobials are accessible across most ICUs in the region, but availability of reserve antibiotics was limited.

The disparities identified in this survey provide priorities for action to improve sepsis outcomes in this region.

## Data Availability

All deidentified survey data is stored on a secure server hosted by The Chinese University of Hong Kong and available on request.

## Appendix 1 Tables

